# Encoding Discordance in the Alzheimer’s Disease A/T/N Framework

**DOI:** 10.64898/2026.07.19.26358425

**Authors:** Lauren Nicole DeLong, Yasamin Salimi, Helena Balabin, Paola Galdi, Jacques D. Fleuriot, Paul Brennan, the Alzheimer’s Disease Neuroimaging Initiative

## Abstract

**INTRODUCTION:** The biomarker-based amyloid/ tau/ neurodegeneration (A/T/N) framework has become a popular staging method for Alzheimer’s disease (AD) research. Previous studies use the framework either as a rule-based or data-driven approach but typically sacrifice either adaptivity or interpretability.

**METHODS:** We present an interpretable, hybrid method, called Neurosymodal Data Fusion, for predicting incident AD in the ADNI dataset. Specifically, we encode the A/T/N framework as a logic program, where the input biomarker features are extracted by one or more neural networks.

**RESULTS:** Our pipeline predicted four-year incident AD with a sensitivity of up to 0.84. Additionally, our models learned scores for each A/T/N profile, denoting relative importances to model predictions. These scores also indicated that empirically-derived cut-off values for the A and T criteria might be uninformative for the ADNI data.

**DISCUSSION:** Our pipeline provides a novel way to use the A/T/N framework that could potentially improve early AD screening years before clinical manifestations.

## 2 Introduction

Although *Alzheimer’s disease (AD)* has no cure yet, early intervention can attenuate symptoms [4,5]. Previous research frameworks to improve our understanding of the disease modelled AD progression in three *syndromal cognitive stages* based on symptoms: (1) *cognitively or clinically unimpaired (CU)*, in which an individual is asymptomatic, (2) *mild cognitive impairment (MCI),* and (3) AD [5–8]. Notably, the MCI stage is particularly heterogeneous: it can indicate pre-clinical AD, but also cognitive impairment due to other causes, in which case a return to the CU stage is possible [9–11]. Thus, many recent studies on AD progression have focused on risk assessment and disease intervention amongst individuals with MCI [12–14].

AD diagnosis is based upon manifestation of clinical symptoms [15], but the underlying AD pathology begins years or decades *before* symptom onset [5,16]. Deposition of the amyloid-β (Aβ) protein and neurofibrillary tangles of the tau protein are two hallmarks of AD [17,18]. These proteinopathies have been detected up to 18 years prior to symptom-based diagnosis [19]. Building on this observation, the *amyloid/ tau/ neurodegeneration (A/T/N) framework* has been proposed as a biomarker-based AD staging scheme [7,15], framing AD as a continuum, in which an individual with a CU or MCI syndromal stage could still have AD pathology [4]. Individuals who are at-risk of AD progression might therefore be identified prior to symptom onset.

The A/T/N framework has been widely adopted [20–22]. However, its discordance with the syndromal cognitive stages complicates the mapping of its biomarker-based stages to clinically meaningful outcomes [15,23]. Additionally, several *different* biomarkers can be used for the A/T/N framework, so biomarker selection may further contribute to heterogeneity *within* staging schemes [20–22] The syndromal MCI stage is particularly heterogeneous [10,11].

Previous efforts have attempted to address these challenges. The *AD Neuroimaging Initiative (ADNI)* [24], a longitudinal cohort study, implemented an additional stage called *early* MCI (EMCI), with milder memory impairment than traditional MCI, thereafter known as the *late* MCI (LMCI) stage. However, it is unclear whether this subclassification is a useful indicator for disease progression [25]. Another study by Ezzati *et al.* [26] explored the use of A/T/N biomarkers to characterize AD progression risk amongst participants with MCI. Specifically, they used several biomarker-based models to distinguish between CU participants and those with AD. Thereafter, they used such models on participants with MCI to classify them as “AD-like” or “CU-like”, indicating whether they are likely to progress to AD or not. However, the models were always either rule-based or data-driven, showing complementary capabilities and weaknesses.

We present a novel pipeline called *Neurosymodal Data Fusion*^1^ for assessing the risk of AD progression, or *incident AD*, based on A/T/N biomarkers. This *Neurosymbolic artificial intelligence (AI)* method [50] combines neural networks (NNs) and deep learning with logical rules, serving as a hybrid between data-driven and rule-based methods. Contrary to the black-box nature of other commonly employed deep learning models for the diagnosis of AD, this hybrid approach enables a far greater degree of interpretability. Our pipeline allows for tracing model predictions back to explicit classification rules that are learned with NNs in a data-driven way, and that describe how a diagnosis label is derived from different types of biomarker evidence. We explored and compared the use of *two* variants of Neurosymodal Data Fusion as well as three baseline models to identify whether individuals diagnosed with MCI would later progress to AD within a four-year time frame.

We found that both Neurosymodal models could identify incident AD amongst MCI participants, outperforming exclusively data-driven models. This indicated that the Neurosymodal models could automatically extract features from images and use them for A/T/N staging in an end-to-end manner. Importantly, by producing probabilities denoting the importance of each A/T/N profile, our approach demonstrates the ways in which uncertainty and discordance surrounding the A/T/N scheme can be accounted for in an interpretable manner.

## 3 Data and Methods

We used A/T/N biomarkers from the ADNI^2^, a longitudinal cohort study designed to investigate AD progression [24]. Within the next sections, we describe the A/T/N framework and the data processing steps taken to ascertain such biomarkers from the ADNI data. Specifically, we introduce our automated and openly available MRI processing pipeline.

Notably, in Sections 3.2.2 and 3.2.5, we describe multiple ways in which the same data was extracted or processed. Since our novel pipeline is a hybrid between data-driven methods and rule-based ones (see Section 3.3), we compared it against two data-driven baseline models, a random forest model [57] and a logistic regression model [49], as well as one rule-based baseline model, a logic program [52] (see Section 3.4.2). Since these models operate differently, they require the same input data in different formats.

Thereafter, we describe the Neurosymodal Data Fusion pipeline and its two model variants in detail. Finally, we discuss the details of model training and evaluation.

### 3.1 The A/T/N Classification Scheme

The A/T/N framework is based upon three criteria: Aβ deposition (A), pathologic tau (T), and neurodegeneration (N) [7,15]. Previous research has identified several clinical and imaging biomarkers to inform each of these three criteria. These biomarkers are derived from various data modalities, including *magnetic resonance images (MRI)*, *positron emission tomography (PET)* scans, and *cerebrospinal fluid (CSF)*.

To use the A/T/N framework, each criterion is binarized as positive (+) or negative (−) according to a chosen biomarker with a pre-determined *cut-off* value or threshold (see Supplementary Table S1). Cut-off values are typically chosen as empirically derived measurements from previous studies (such as those described in Section 3.2.2). Following the binarization of each of the three criteria, there are eight resultant profiles, summarized in Table 1, which can be interpreted as belonging to three over-arching categories [15].

**Table 1.** A/T/N profiles and their pathological categories, adapted from Jack *et al.*, 2014 [15].

| A/T/N Profile | Description | Category |
| --- | --- | --- |
| A-/T-/N- | Normal biomarkers | Normal |
| A+/T-/N- | AD pathology | AD continuum |
| A+/T+/N- | AD | AD continuum |
| A+/T+/N+ | AD | AD continuum |
| A+/T-/N+ | AD pathology | AD continuum |
| A-/T+/N- | Non-AD pathologic change | Non-AD pathology |
| A-/T-/N+ | Non-AD pathologic change | Non-AD pathology |
| A-/T+/N+ | Non-AD pathologic change | Non-AD pathology |

Firstly, if a participant has no positive criteria (*i.e.,* A−/T−/N−), they are said to have normal biomarkers and therefore no detectable disease pathology. Next, a participant is on the *AD continuum* and shows AD *pathology* if they have A *positivity* (*i.e.,* A+/T±/N±). Within the AD continuum, AD itself is defined by *both* A *and* T positivity (A+/T+/N±) since Aβ and tau proteinopathies are hallmarks of the disease [17,18]. Neurodegeneration, on the other hand, is nonspecific to AD [15], so all other profiles are categorized as *non-AD pathology*.

### 3.2 ADNI Dataset

The ADNI was launched in 2003 as a public-private partnership, comprising several numbered phases, ADNI 1 to ADNI 4, which is currently ongoing, as well as ADNI-GO, which focused primarily on biomarkers for early disease detection [24]. The various phases of ADNI were tailored for specific research goals. For example, while ADNI 1 was focused on testing whether MRI, PET, and neuropsychological assessments can be combined to measure the progression of MCI and early AD, ADNI 4 is focused on validating biomarkers and improving the generalizability of AD research by increasing participant diversity [24].

Since the ADNI is a longitudinal study, participants typically return for data collection over several subsequent visits every six to twelve months. Because some participants miss visits or drop out of the study, the number of visits and the availability of biomarker measurements varies by participant [30]. Additionally, participants from earlier stages often rollover into subsequent phases. At the date of access (October 24, 2024), the ADNI contained longitudinal data on 5,396 participants aged 55-90 years at baseline. This data was collected across 105 centres within the United States and Canada. Of available data, ADNI contains both raw and processed versions of MRIs and PET scans, CSF biomarker information, and more [30,31].

#### 3.2.1 MRI Processing

Our proposed Neurosymodal models require MRIs as input to automatically determine N positivity in the A/T/N framework. Therefore, pre-processed T1-weighted MRI scans from ADNI were downloaded for 1,193 participants at every available visit. Pre-processing steps included N3 bias-field corrections [32] as well as gradient non-linearity corrections [33], where available [24].

Our MRI pre-processing pipeline (see Supplementary Figure S1), includes several additional steps. First, we skullstripped MRIs with the FMRIB Software Library (FSL) [34] implemented in Nipype [35]. Following skullstripping, we extracted the two-dimensional (2D) images comprising the center, axial slices of each MRI. Finally, MRIs were downsampled to 150×150 pixels.

#### 3.2.2 A/T/N Biomarkers

To determine A and T positivity (A+ and T+, respectively), we extracted biomarkers from the CSF and PET uptake data, as described in Bucci *et al.* [20]. In summary, CSF data was previously collected via lumbar punctures, and it provides information about A+ based on levels of amyloid-β1-42 (Aβ1-42) (cut-off ≤880 picograms per milliliter (pg/mL)) and T+ based on levels of phosphorylated Tau 181 (pTau) (cut-off ≥26.64 pg/mL). PET uptake values describe functional information which is captured through radioactive tracers [28]. From PET uptake values, we ascertained two A+ biomarkers: the standardized uptake value ratios (SUVR) for ^18^F-florbetapir (FBP) and ^18^F-florbetaben (FBB), two tracers that detect amyloid plaques within the brain [28]. The cut-offs for FBP and FBB were ≥ 1.11 and 1.08 SUVR, respectively, which were previously validated on ADNI data [16,36,37]. Finally, another tracer, ^18^F-labeled tau PET ligand flortaucipir (FTP) shows Tau deposition in the brain [28], so it is useful for assessing three additional T+ biomarkers. Previous studies identified three biomarkers regarding specific regions of interest (ROIs) over composite regions of the brain. These are referred to as T1, T2, and T3 ROIs, and cover a temporal meta-ROI, the inferior temporal cortex, and entorhinal cortex, respectively. T+ cut-offs for these ROIs were ≥ 1.37, 1.31, and 1.39 SUVR, respectively [20,38,39].

In contrast to the Neurosymodal pipeline, which can determine N positivity directly from MRIs, our three baseline models require ground-truth N+ biomarker values. Previous studies found two MRI-derived N+ biomarkers using FreeSurfer [40] analyses: (1) hippocampal volume, adjusted for intracranial volume (HVa) [41] and (2) temporal lobe cortical thickness (JCT), involving a composite region between the entorhinal, inferior temporal, middle temporal, and fusiform ROIs. Cut-off values of *<*6723 cubic millimeters (mm^3^) [42] and *<*2.67 millimeters (mm) [43] were previously described for these values, respectively. These values were computed according to Jack *et al.* [41], but we also adjusted HVa for FreeSurfer software version (v. 4.3, 5.1, 6.0) to account for discrepancies between versions [44]. Finally, total Tau (tTau) is a third, CSF-based N+ biomarker, and we used its established cut-off of ≥ 300 pg/mL [20,45]. We summarize all biomarkers and their respective cut-off values within Supplementary Table S1, and list all required files for obtaining these values in Supplementary Table S2.

#### 3.2.3 Data Point Construction

As mentioned in Section 2, Neurosymodal Data Fusion combines deep learning with logical rules. Since deep learning architectures typically require a large number of samples for training [46,47], we maximized sample size by using data across *all* available visits. Henceforth, we refer to each sample as a *data point*, defined here as an MRI (from Section 3.2.1) with corresponding PET and/or CSF-derived A and T biomarkers (from Section 3.2.2). Thus, several data points could represent the disease stages of the *same* participant at *different* visits. We matched each MRI to its corresponding A/T biomarkers according to whether (1) the records belong to the same participant, and (2) the CSF or PET data for A/T was acquired within ±13 months (395 days) of the MRI acquisition date, as is consistent with previous studies [20] (Supplementary Figure S2). If an MRI had no biomarker data within that time frame, it was excluded (Supplementary Figure S3).

We assigned AD, MCI and CU labels for each data point based on the participant’s diagnosis on the date of MRI acquisition. Additionally, we ascertained labels denoting *incident* AD for each of the MCI-stage data points. We derived such labels from subsequent visit information within the four-year time frame following each data point’s *latest* examination date. In other words, if the data point’s latest biomarker was taken 395 days following MRI acquisition, the four years following that date were considered. We selected a duration of four years because it is the common duration of AD clinical prevention trials [12,26]. We labelled those who had transitioned to AD as belonging to the positive class, most analogous to “AD-like”, and those who did not to the negative class, most analogous to the “CU-like” class. To account for dropout, MCI data points with no subsequent visit data within that time frame were excluded (Supplementary Figure S3). Additionally, using a two-sided Mann-Whitney U test (two-sided, α=0.05) [48], we confirmed that follow-up durations were not significantly different between these two classes (*p* =0.30).

#### 3.2.4 Extracted Dataset and Biomarker Availability

Following MRI processing, biomarker matching, and dropout-based exclusions, our resulting dataset contained 3,743 data points from 964 participants (Supplementary Figure S3). Of these, nearly all data points (99.9%; 100.0% of participants) had at least one biomarker informing the A criterion, 79.4% (83.9% of participants) had at least one biomarker informing the T criterion, and 89.4% (97.7% of participants) had at least one biomarker informing the N criterion (Supplementary Table S3). Although each data point contained an MRI, the MRI-derived FreeSurfer outputs (described in Section 3.2.2), were not always available.

Figure 1 shows both relative biomarker availability and the frequency for each combination of biomarkers amongst these 3,743 data points. Overall, biomarkers derived from CSF tended to have better availability than those derived from imaging modalities. One exception to this pattern is the A biomarker, FBP, which was available for 98.0% of data points. Aside from FBP, Aβ1-42 was the next most available A biomarker (77.9% of data points). Of the four possible T biomarkers, pTau was most available (77.9% of data points). Finally, of the N biomarkers, tTau was also most available at the same rate (77.9% of data points). HVa and JCT, two MRI-derived N biomarkers, were available for 32.0% and 50.7% of data points. Other biomarkers, including FBB for A and the three PET-based ROIs for T, were available for very few data points (0.2%, 2.9%, 2.9%, and 2.9%, respectively).

**Figure 1.**
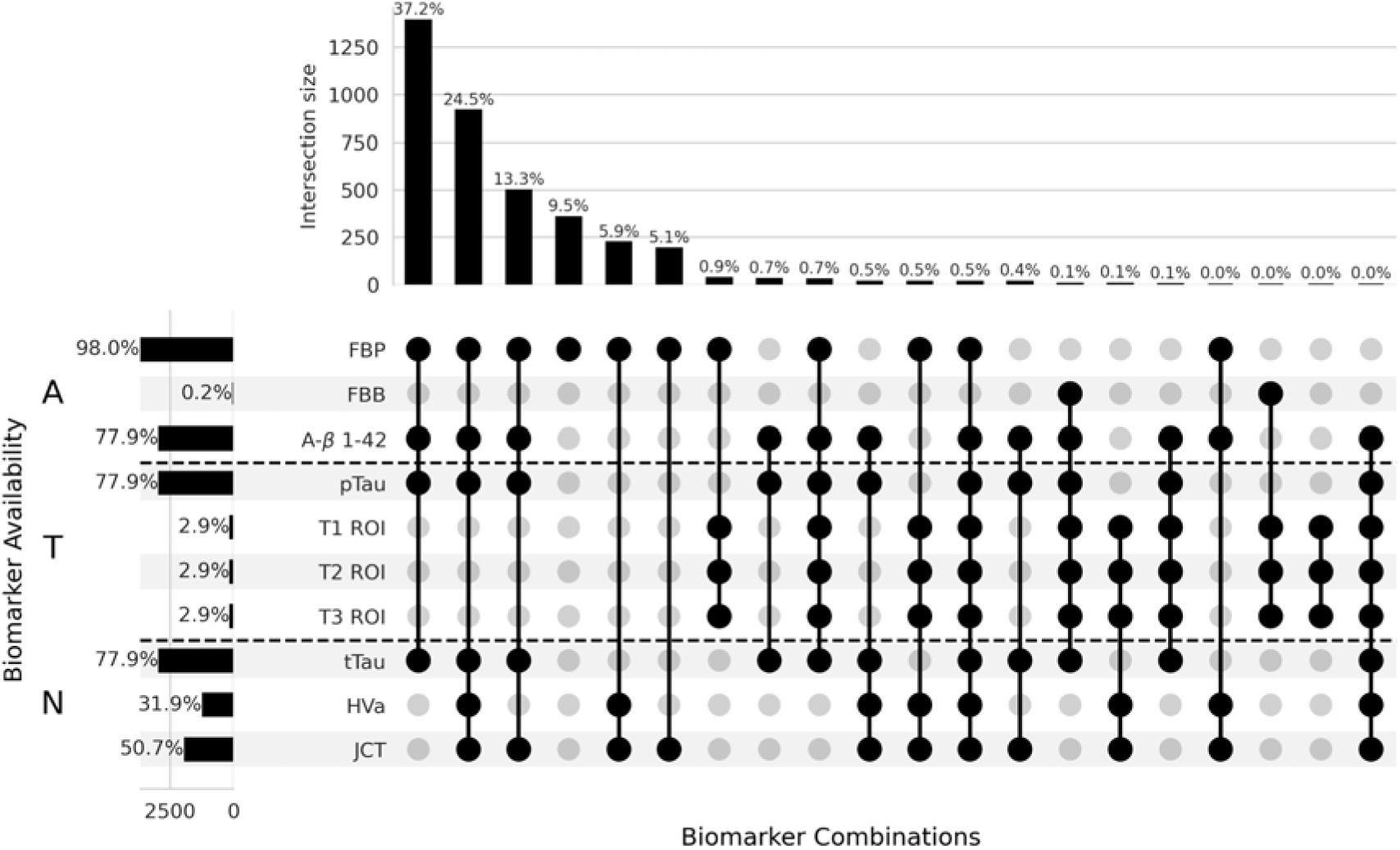
A/T/N Biomarker availability and combinations for the 3,743 data points used. For instance, the first column represents the 37.2% of data points for which FBP, Aβ1-42, pTau and tTau measurements were available.

#### 3.2.5 Biomarker Input Values

As we will discuss in Section 3.3., the Neurosymodal Data Fusion pipeline is modular, so there are various ways to use it, dependent upon data availability. Additionally, the two Neurosymodal models as well the baseline models (Section 3.4.2) require input data in various formats. Specifically, while some require the binarization of certain biomarker values (see Section 3.1) *beforehand,* others require continuous biomarker values as input, determining such thresholds intrinsically. Therefore, aside from MRI processing described in Section 3.2.1, we prepared biomarker data from Section 3.2.2 in two different ways.

Some models, including one of the Neurosymodal models (Section 3.3.1), the random forest model, and the logistic regression model, require continuous biomarker values as input. In such cases, it was important to consider data availability (see Section 3.2.4). Firstly, we excluded values for FBB and the T1, T2, and T3 ROIs due to insufficient data availability. Of the included biomarkers, we imputed any missing values with fixed, or constant, values. Specifically, we used each biomarker’s respective cut-off value, as specified in Supplementary Table S1, since cut-off values represent neutral or borderline pathology. Finally, according to previously established best practices [49], we scaled continuous biomarker values between zero and one based on minimum and maximum values in the training set, which is introduced in the next section.

Other models, including the other Neurosymodal model and the baseline logic program, require the *probabilities* of A+, T+, and N+ as input. We compute these as the *concordances* between available A, T, and N biomarkers. Using the empirically-derived cut-off values described in Section 3.2.2, we determined, in a rule-based manner, the positivity of each data point’s biomarkers through the binarization technique described in Section 3.1. From this information, we computed the concordances as the proportions of *positive* biomarkers to the *total available* biomarkers for each criterion and participant. For example, if two of three A biomarkers were available for a participant, and one of these two showed positivity, the participant would be assigned a probability of 0.5, for A+. We show the distributions of these concordances, per criterion, in Supplementary Figure S4a, S4b, and S4c, respectively. Notably, all three distributions are tri-modal: each has modes at 0.0, 1.0, and a third mode in between. Where *no* biomarkers were available for a criterion, missing values were imputed as the central mode of each criterion’s concordance distribution (0.25, 0.50, and 0.50 for A, T, and N, respectively).

#### 3.2.6 Training, Validation, Test, and MCI Sets

We split the extracted dataset into *four* disjoint sets. To create the first three, we divided 1,175 CU-stage data points (from 283 participants) and 492 AD-stage data points (from 148 participants) into stratified *training*, *validation*, and *test sets* based on a 70%/15%/15% split. We used the training set, which comprised 1,154 data points (816 CU, 338 AD), to fit model parameters, and we used the validation set, which comprised 259 data points (182 CU, 77 AD), to optimize model hyperparameters (see Appendix A). Finally, we used the test set, comprising 254 data points (177 CU, 77 AD), to gauge whether a model could distinguish between AD and CU stages amongst a set of previously unseen data points (Table 2).

**Table 2.** Syndromal stage distribution across training (*n*=1,154 participants), validation (*n*=259 participants), test (*n*=254 participants), and MCI (*n*=2,074 participants) sets. *The syndromal stage distribution across the MCI set should be understood as those who did *not* experience incident AD (“CU-like”) and those who did (“AD-like”).

| Syndromal Stage | Training |  | Validation |  | Testing |  | MCI* |  |
| --- | --- | --- | --- | --- | --- | --- | --- | --- |
|  | Count | Ratio | Count | Ratio | Count | Ratio | Count | Ratio |
| CU | 816 | 0.71 | 182 | 0.70 | 177 | 0.70 | 1,349 | 0.65 |
| AD | 338 | 0.29 | 77 | 0.30 | 77 | 0.30 | 725 | 0.35 |

We created a *fourth* set, called the *MCI set*, comprising previously unseen data points exclusively within the MCI stage. We designed the MCI set to determine whether models that were trained to distinguish between AD and CU stages could subsequently predict whether participants in the MCI set would transition to the AD stage (*i.e.*, a data point is predicted as “AD-like”) or not (*i.e.*, a data point is predicted as “CU-like”). Although models *could*, in theory, be trained directly on the MCI set, rather than the training set, labels denoting progression would be unavailable for those with MCI in the *present*. By training models *first* to distinguish between AD and CU stages and then, subsequently, evaluating on data points in the MCI stage, our study shows the extent to which models can generalize to predicting future or *incident* AD. Specifically, the MCI set involved 2,074 data points (from 533 participants). Of these, 725 data points (from 189 participants) were in the positive class (transitioned to AD), 1,349 data points (from 344 participants) were in the negative class.

### 3.3 Neurosymodal Data Fusion

*Neurosymodal Data Fusion*^3^, was designed and tested for assessing the risk of incident AD, based on A/T/N biomarkers. It consists of two modules: a neural module, comprising at least one NN, and a symbolic module, involving a probabilistic logic program [52]. As input, both models take MRIs with all available A and T biomarker data for a set of participants. In both cases, the neural module extracts information about whether participants show A, T, or N positivity. The second, symbolic module combines the output of the neural module to determine the probabilities, for each participant, of being assigned to “CU-like” or “AD-like” classes.

Importantly, the pipeline’s modular design allows it to be customized according to a study’s needs. To demonstrate this, two variants, or Neurosymodal models, were created with different versions of the neural module. **Neurosymodal model A**, as shown in Figure 2, involves one NN, whereas **Neurosymodal model B**, shown in Figure 3, uses three NNs. In short, the two models differ through the ways in which the A and T criteria are represented. While both models use multiple available biomarkers, model A utilizes the concordances mentioned in Section 3.2.5, and model B learns the most appropriate cut-offs through its two additionalNNs. Further details on the structure and implementation of the Neurosymodal pipeline as well as the two model variants are discussed in the next sections.

**Figure 2.**
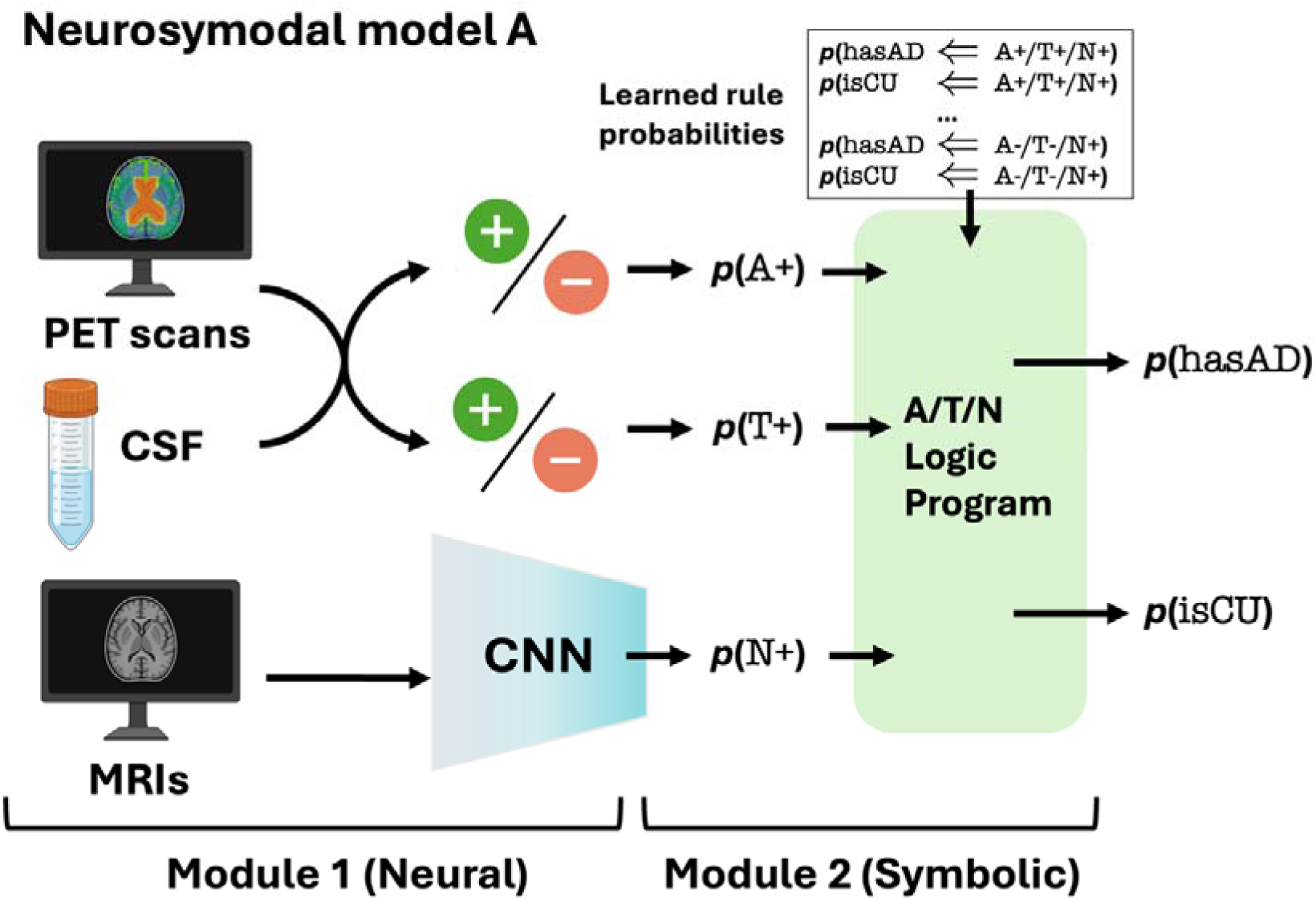
Neurosymodal model. **A.** This Neurosymodal model starts with a convolutional neural network (CNN) which extracts information about neurodegeneration (*p*(N+)) from MRIs. The probabilities of A+ and T+ (*p*(A+), *p*(T+)) are computed as the concordances of A and T biomarkers. Thereafter, these probabilities are fed into a second, symbolic module comprising a probabilistic logic program. In addition to the probabilities of each criterion, rule probabilities, which are learned by the model, are provided to the symbolic module. Using logical inference, the second module determines the probability that each data point is “CU-like” (*p*(isCU)) or “AD-like” (*p*(hasAD)). Partially created in https://BioRender.com.

**Figure 3.**
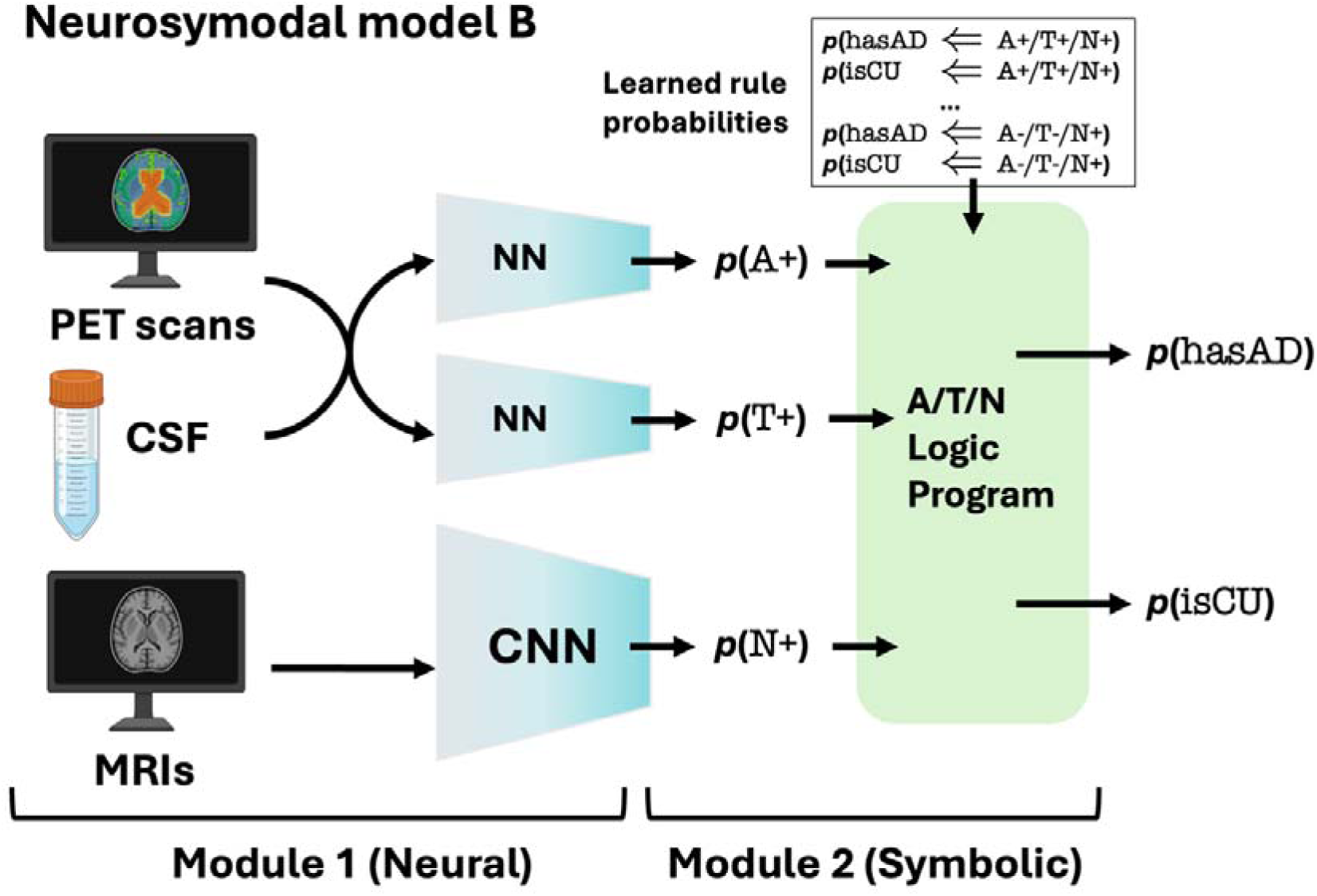
Neurosymodal model. **B.** This Neurosymodal model starts with three NNs, one of which is a CNN that extracts information about neurodegeneration (*p*(N+)) from MRIs. The probabilities of A+ and T+ (*p*(A+), *p*(T+)) are determined by two single-layer NNs on A and T biomarkers, respectively. Thereafter, these probabilities are fed into a second, symbolic module comprising a probabilistic logic program. In addition to the probabilities of each criterion, rule probabilities, which are learned by the model, are provided to the symbolic module. Using logical inference, the second module determines the probability that each data point is “CU-like” (*p*(isCU)) or “AD-like” (*p*(hasAD)). Partially created in https://BioRender.com.

#### 3.3.1 Module 1: The Neural Module

The neural module of the pipeline (see left half of Figure 2 and 3) takes biomarker data as input and assigns the probabilities, for each participant, of having A, T, and N positivity. Since different AD datasets have varying levels of biomarker availability [30] or known cut-off values [37,53], methods to determine A, T, and N positivity can vary by study. Therefore, the neural module can be customized. Neurosymodal models A and B demonstrate two ways to do this.

Both models have neural modules involving a CNN (Appendix B) which takes structural MRIs as input and predicts the probability of neurodegeneration, or N+, from each image. These probabilities serve as input into the second module, and the output of the second module is used to update the parameters of the CNN. Neurosymodal model B also involves two additional, single-layer NNs within its neural module: one takes the scaled FBP and Aβ1-42 values to predict the probability of A+, while the other does the same on scaled pTau values for T+. All three NNs in Neurosymodal model B were trained in tandem through methods described in Section 3.4. In contrast, Neurosymodal model A uses the concordances from Section 3.2.5 as the probabilities of A+ and T+.

#### 3.3.2 Module 2: The Symbolic Module

For both Neurosymodal models, their second, symbolic module (see right halves of Figure 2 and 3) involves a *probabilistic logic program* encoding the A/T/N framework. Logic programs represent knowledge through rules and facts. Facts describe aspects that are known to be true. For example, “*Participant X shows Amyloid pathology (A+)*” is a fact which could be derived from biomarker data. As mentioned, there is discordance within the A/T/N framework with respect to biomarker selection. Thus, there is some uncertainty surrounding this fact. Using *probabilistic* logic, we can represent this fact and its uncertainty as the following, in which 0.25 represents a probability, or degree of certainty, that *Participant X* has A positivity:

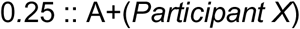

Ultimately, the logic program is grounded in probabilistic facts which represent the probability that each participant has A, T, or N positivity. The probabilities come from the NNs in module 1.

*Rules*, the other key component to logic programs, describe patterns which can be used to deduce new facts. In a probabilistic logic program, rules may also have associated probabilities. For instance, the following rule says that a participant with A, T, and N positivity has a 0.95 probability of AD:

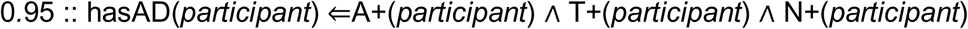

The *predicates*, which are written in the form, predicate(*participant*), can describe situations in which a participant has AD as well as A, T, and N positivity, respectively. We list all predicates in the Neurosymodal logic program and their interpretations in Supplementary Table S4. The implication arrow (⇐) separates the rule *head* from the rule *body*. If the body is evaluated to be true, then the rule is said to be *satisfied*, and the head is also deemed true. In other words, the head is *entailed* by the body. Finally, the ∧ symbol is a logical “and”, an operator which, in this case, specifies that *all* predicates in the body should be true for the rule to be satisfied.

Using logical facts and rules, we formally represent the A/T/N scheme by encoding each of the eight A/T/N profiles in Supplementary Table S5 as the bodies of *two* logical rules: one which entails that a participant has AD and another which entails that a participant is CU. This accounts for the idea that AD and CU participants can belong to any of the eight A/T/N profiles. Therefore, our logic program comprised sixteen rules with corresponding rule probabilities between zero and one, inclusive. However, since participants must be classified into one of the two classes, the probabilities of a participant having AD, *p*(AD), and of being CU, *p*(CU), must sum to one for each rule body. In other words, for each rule body:

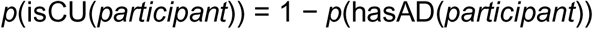

Importantly, the Neurosymodal Data Fusion pipeline allows these rule probabilities to be *learned* as the neural module is trained. Once learned, the probabilities represent the relative importance of each rule body for predicting the AD and CU classes. Using logical facts and rules, the symbolic module formally represents the A/T/N framework. Importantly, by using *probabilistic* logic, we encode uncertainty in two key ways. Firstly, probabilistic *facts* account for any uncertainty surrounding biomarker data, such as that due to discordance between biomarkers of the same criterion. Secondly, probabilistic *rules* account for the discordance between the A/T/N and syndromal cognitive staging schemes.

### 3.4 Training and Evaluation

In the next subsection, we discuss the details around model training and evaluation. Put succinctly, we trained or fit models to distinguish between AD and CU participants. Thereafter, we used the trained models to assess the risk of incident AD within four years by classifying MCI participants into “CU-like” or “AD-like” groups. To assess predictive performance, we compared the Neurosymodal models to both rule-based and data-driven baseline models. Additionally, we analyzed the alignment of predicted classifications against the EMCI and LMCI subclasses of the MCI stage.

#### 3.4.1 Model Implementation and Training

We implemented the Neurosymodal pipeline in an end-to-end manner (see Appendix C). Notably, we designed it so that, during training, the three NNs in Neurosymodal model B could be updated simultaneously. We trained both Neurosymodal models on the training set, representing individuals who are either in the CU or AD stages. To account for label imbalance, we weighted the AD class in the loss function by 2.4 times more than the CU class (*i.e.*, the relative proportion of CU versus AD data points).

#### 3.4.2 Model Evaluation

Following training, we evaluated model performances on two held-out datasets. First, we evaluated each model on the test set to assess its capability to distinguish previously unseen CU and AD data points. Then, as mentioned in Section 3.2.6, we used the MCI set to assess whether each model could predict incident AD amongst MCI-stage data points.

We measured model performances using *sensitivity*, a model’s ability to identify the positive class (“AD-like”), *specificity*, a model’s ability to identify the negative class (“CU-like”) [54], and *macro F1 score*, a measure of a model’s overall predictive capabilities between the two classes [55,56].

Finally, we compared model performances to three baseline models. The first of such baseline models was a probabilistic logic program (module 2) *without* any NN. As input, the logic program took A/T/N probabilities based on concordances (Section 3.2.5). The other two baseline models included two interpretable, data-driven models based on (1) a random forest [57] and (2) logistic regression [49]. For both data-driven models, we used the *continuous, scaled* biomarker values. Additionally, for data-driven models, weighted class labels were as described in Section 3.4.1. Appendix A describes hyperparameter optimization for each model.

#### 3.4.3 Alignment with Early and Late-stage MCI

To analyze how closely model predictions aligned with existing domain knowledge, we compared the predictions on the MCI set in a contingency table [58] against each data point’s respective EMCI and LMCI labels (see Section 3.2.6). Given that the LMCI classification was designed to denote a more severe or further form of progression toward the AD stage than EMCI, one might expect that the model’s predictions toward “CU-like” and “AD-like” would align with the EMCI and LMCI subclassifications, respectively. To statistically test this concept, we used Fisher’s Exact Test (one-sided, α =0.05) [59].

## 4 Results

### 4.1 Prediction of AD versus CU

All five models yielded high predictive performance, with macro F1 scores between 0.71-0.78. This indicates that A/T/N biomarkers are useful for distinguishing the AD stage from the CU stage. Between the five models, the random forest model had the best F1 score (0.78), followed by Neurosymodal model B and the logistic regression (Log. Reg.) model (0.76). Both the random forest and logistic regression models had the best specificity (0.85) on the test set, but the lowest sensitivities (0.74 and 0.66). Neurosymodal model A, on the other hand, had the highest sensitivity (0.84) but a lower specificity (0.67). Neurosymodal model B had a slightly lower sensitivity than Neurosymodal model A (0.79) but a better specificity (0.77). In summary, the random forest and logistic regression models were best at predicting the CU class, the Neurosymodal model A was best at predicting the AD class, and Neurosymodal model B struck a balance between the two.

### 4.2 Prediction of Incident AD

Of the five models, the random forest model had the highest macro F1 score (0.74), closely followed by the logistic regression model (0.73), then Neurosymodal model B (0.72). While the random forest and logistic regression models also had the best specificity (both 0.78), the logic program and Neurosymodal model A had the best sensitivity (0.81), followed by Neurosymodal model B (0.75). Higher sensitivities indicate that the Neurosymodal models and logic program, all of which encode the A/T/N profiles, were better at predicting the positive class, or incident AD, than the data-driven models. Additionally, as in the previous section (Section 4.1), Neurosymodal model B achieved the most balanced sensitivity and specificity (0.75 and 0.72, respectively) compared to the other models.

Although the five models showed varying capabilities, all models had high performance metrics for both predictive tasks. Thus, the A/T/N biomarkers are informative for predicting both current and incident AD. This notion is also supported by the rule probabilities reported in the next section.

### 4.3 Rule Probabilities for Interpretability

Both the Neurosymodal models learn rule probabilities for each of the A/T/N profiles. These rule probabilities can serve as measures of how important each A/T/N profile is to the “AD-like” (hasAD(*participant*)) or “CU-like” (isCU(*participant*)) classifications (Table 4).

**Table 3.** (Top) Performance metrics for distinguishing between AD (*n*=77 participants) and CU (*n*=177 participants) data points in the test set (*n*=254 participants). (Bottom) Performance metrics for predicting incident AD (*n*=725 participants) in the MCI set (*n*=2,074 participants). RF=random forest model; Log. Reg.=logistic regression model. Best metrics are in bold.

| Test set metrics |  |  |  |  |  |
| --- | --- | --- | --- | --- | --- |
|  | Neurosymodal model A | Neurosymodal model B | Logic Program | RF | Log. Reg. |
| Macro F1 | 0.71 | 0.76 | 0.71 | **0.78** | 0.76 |
| Sensitivity | **0.84** | 0.79 | 0.75 | 0.74 | 0.66 |
| Specificity | 0.67 | 0.77 | 0.72 | **0.85** | **0.85** |
| MCI set metrics |  |  |  |  |  |
|  | Neurosymodal model A | Neurosymodal model B | Logic Program | RF | Log. Reg. |
| Macro F1 | 0.69 | 0.72 | 0.70 | **0.74** | 0.73 |
| Sensitivity | **0.81** | 0.75 | **0.81** | 0.70 | 0.69 |
| Specificity | 0.63 | 0.72 | 0.65 | **0.78** | **0.78** |

**Table 4.** Learned rule probabilities for the Neurosymodal models.

| A/T/N Category | hasAD(*participant*) |  | isCU(*participant*) |  | Rule Body | A/T/N Profile |
| --- | --- | --- | --- | --- | --- | --- |
|  | Neurosymodal model A | Neurosymodal model B | Neurosymodal model A | Neurosymodal model B |  |  |
| Normal biomarkers | <0.01 | <0.01 | >0.99 | >0.99 | $A-(participant) \wedge T-(participant) \wedge N-(participant)$ | A-/T-/N- |
| AD continuum | 0.06 | >0.99 | 0.94 | <0.01 | $A+(participant) \wedge T-(participant) \wedge N-(participant)$ | A+/T-/N- |
| | >0.99 | 0.96 | <0.01 | 0.04 | $A+(participant) \wedge T+(participant) \wedge N-(participant)$ | A+/T+/N- |
| | 0.44 | 0.43 | 0.56 | 0.57 | $A+(participant) \wedge T+(participant) \wedge N+(participant)$ | A+/T+/N+ |
| | 0.79 | 0.60 | 0.21 | 0.40 | $A+(participant) \wedge T-(participant) \wedge N+(participant)$ | A+/T-/N+ |
| Non-AD pathology | <0.01 | <0.01 | >0.99 | >0.99 | $A-(participant) \wedge T+(participant) \wedge N-(participant)$ | A-/T+/N- |
| | 0.33 | 0.23 | 0.67 | 0.77 | $A-(participant) \wedge T-(participant) \wedge N+(participant)$ | A-/T-/N+ |
| | 0.12 | 0.86 | 0.88 | 0.14 | $A-(participant) \wedge T+(participant) \wedge N+(participant)$ | A-/T+/N+ |

There are some clear consistencies between the two models. Firstly, the profile representing normal biomarkers, A−/T−/N−, had high probabilities for the isCU(*participant*) rule head (*>*0.99 for both models). This indicates that the A−/T−/N− profile is consistently more important to the “CU-like” class than the “AD-like” one, an idea which is consistent with the original A/T/N framework [7]. Additionally, the A+/T+/N− profile had high probabilities for the hasAD(*participant*) rule head (*>*0.99 and 0.96 for models A and B, respectively), indicating an importance for predicting the “AD-like” class. This also aligns with the idea that the proteinopathies represented by the A and T criteria, Aβ plaques and tau tangles, are hallmarks of AD [17,18].

There are also some clear differences between the rule probabilities for the two models. Firstly, Neurosymodal B associated the A−/T+/N+ with a high probability for the hasAD(*participant*) rule head (0.86), whereas Neurosymodal model A gave it a low probability (0.12). Additionally, the opposite, A+/T−/N− profile had a low probability for the hasAD(*participant*) rule head under Neurosymodal model A (0.06) but a very high probability under Neurosymodal model B (*>*0.99). These results could indicate that one or more of the empirically-derived cut-off values for A+ or T+ used in Neurosymodal model A were uninformative or biased by the participant data on which they were derived, an idea which has been discussed in previous literature [53,60].

### 4.4 Early and Late-Stage Classifications

We compared each Neurosymodal model’s predictions on the MCI set to the respective EMCI and LMCI subclasses in the ADNI data. While previous research has called for further validation of these subclasses [25], they can serve as a source of additional domain knowledge to which prediction results can be compared. Intuitively, one would predict that the “CU-like” predictions would correspond with the EMCI subclass, as both indicate earlier stages or a lack of progression, whereas the “AD-like” predictions would correspond with the LMCI subclass. This comparison is shown in Table 5.

**Table 5.** Overlaps between Early and Late-stage subclasses of MCI (EMCI and LMCI, respectively) and the Neurosymodal model A predictions (top) as well as Neurosymodal B predictions (bottom) for incident AD in the MCI set.

|  | “CU-like” | “AD-like” |
| --- | --- | --- |
|  | Neurosymodal model A |  |
| **EMCI** | 586 | 479 |
| **LMCI** | 407 | 604 |
|  | Neurosymodal model B |  |
| **EMCI** | 680 | 383 |
| **LMCI** | 464 | 547 |

Of the data points in the MCI set classified as “CU-like” by model A, over half (59.0% or 586 data points) were labeled as EMCI. This number was higher for model B, at 64.0% or 680 data points. Of the data points in the MCI set classified as “AD-like” by model A, 55.8% or 604 data points were labeled as LMCI. This number was slightly lower for model B, at 54.1% or 547 data points. Based on Fisher’s Exact Test, there was significant alignment for both models (*p* = 1.03×10^−11^ and *p* = 7.95×10^−17^ for model A results and model B results, respectively). Nevertheless, the relevance of EMCI and LMCI subclasses to this task requires further investigation.

## 5 Discussion

We explored the use of two novel, neurosymbolic models, based upon our Neurosymodal Data Fusion pipeline, for predicting the subsequent progression to AD from the MCI stage. The Neurosymodal Data Fusion pipeline serves as a novel way to use the A/T/N framework in a hybrid fashion between data-driven approaches and rule-based approaches. Specifically, by encoding discordance through probabilistic logic, it was demonstrated that the A/T/N framework can be useful for predicting incident AD. Furthermore, the rule probabilities learned by a Neurosymodal model provide several additional benefits. Firstly, they are interpretable to the end-user: they help to explain which A/T/N profiles were most important to “CU-like” versus “AD-like” classifications. Additionally, they can help to identify potential biases in the training dataset, especially when the probabilities do not necessarily align with expectations. Moreover, the proposed neurosymbolic models yielded a higher sensitivity (up to 0.84, see Table 3) compared to the baseline models, potentially benefitting screening tools for early AD detection. Finally, these rule probabilities can be tailored via re-training to better fit and represent other datasets.

Notably, compared to other cohort datasets, ADNI has relatively complete A/T/N biomarker data [30]. Furthermore, ADNI has been used in thousands of published studies [61], so empirical cut-off values have been extensively researched and validated [37,53]. However, other studies may involve more uncertainty around the A/T/N framework due to varying data availability [30] and lack of validation [62]. In such cases, data-driven approaches may be necessary to determine one or more of the A/T/N criteria. The Neurosymodal pipeline provides a way in which the A/T/N framework can be used along with data-driven methods, like the CNN for N+ determination. Therefore, Neurosymodal Data Fusion can be customized to use any combination of cut-off-based or data-driven methods for determining A, T, and N criteria.

### 5.1 Related Work

The predictive tasks in this study were similar to those in Ezzati *et al.* [26]. However, our study represented the A/T/N framework using a novel neurosymbolic approach. Specifically, we introduced ways to represent uncertainty within the A/T/N scheme through probabilistic rules and facts. Consequently, we found that, in contrast to Ezzati *et al.*, our logic program and Neurosymodal models could better predict incident AD than data-driven models alone.

Recently, two studies have used neurosymbolic approaches within the context of AD. The first also proposes a neurosymbolic approach centered around probabilistic logic programming [63]. However, it is used to predict cognitive decline scores, rather than transition to MCI. The second, called NeuroSymAD [64], combines a NN over MRIs with logical rules like we do. Rather than focusing on the capabilities of the A/T/N scheme, however, its logic program is derived from a large language model that extracts rules from clinically relevant literature. Additionally, NeuroSymAD is used to predict AD diagnosis rather than progression.

### 5.2 Limitations and Prospective Directions

Previous studies have shown that AD progression patterns differ across study cohorts [9,65], so a key prospective direction includes the validation of the study results upon another AD dataset. Other prospective directions for research could involve the inclusion of other model features, such as demographic variables or speech patterns [66], as well as accounting for the longitudinal nature of the data. One key limitation of the Neurosymodal models is the treatment of data at each visit as independent data points, thereby disregarding any temporal aspects of the data. Additionally, although it was ensured that there was no crossover between sets with respect to participants, there were likely dependencies between data points belonging to the same participant. Thus, another prospective direction is the use of a pre-trained CNN within the neural module of the Neurosymodal pipeline. In other words, one could incorporate a CNN which was trained on a similar but different task and dataset. Consequently, this would reduce the need for a larger dataset [67].

## 6 Conclusions

We presented the novel Neurosymodal Data Fusion pipeline. Using this pipeline, we trained two Neurosymodal models to predict incident AD, achieving higher sensitivities to AD progression within four years than exclusively data-driven ones. Our study showed that a neurosymbolic paradigm can be useful for encoding uncertainty within structured domain knowledge. Specifically, the Neurosymodal Data Fusion pipeline serves as a unique way to represent and use the A/T/N framework, a rule-based system designed by AD researchers. Importantly, it can use a mixture of data-driven methods, such as the CNN for N+ determination, or domain-specific knowledge, such as empirically-derived cut-off values, to make predictions. This enables customization of the pipeline to fit the needs of a particular study.

## Data Availability

Data were obtained from the ADNI database (adni.loni.usc.edu).

https://adni.loni.usc.edu/

## Consent Statement

Data were obtained from the ADNI database (adni.loni.usc.edu). All ADNI participants have provided informed consent.

## Footnotes

1 https://github.com/laurendelong21/neurosymodal

2 adni.loni.usc.edu

3 https://github.com/laurendelong21/neurosymodal

## References

[1] Scheltens P, De Strooper B, Kivipelto M, Holstege H, Chételat G, Teunissen CE, et al. Alzheimer’s disease. The Lancet 2021;397:1577–90.

[2] Better MA. Alzheimer’s disease facts and figures. Alzheimers Dement 2023;19:1598–695.

[3] Aranda MP, Kremer IN, Hinton L, Zissimopoulos J, Whitmer RA, Hummel CH, et al. Impact of dementia: Health disparities, population trends, care interventions, and economic costs. J Am Geriatr Soc 2021;69:1774–83.

[4] Aisen PS, Cummings J, Jack CR, Morris JC, Sperling R, Frölich L, et al. On the path to 2025: understanding the Alzheimer’s disease continuum. Alzheimers Res Ther 2017;9:1–10.

[5] Sperling RA, Aisen PS, Beckett LA, Bennett DA, Craft S, Fagan AM, et al. Toward defining the preclinical stages of Alzheimer’s disease: Recommendations from the National Institute on Aging-Alzheimer’s Association workgroups on diagnostic guidelines for Alzheimer’s disease. Alzheimer’s & Dementia 2011;7:280–92.

[6] McKhann GM, Knopman DS, Chertkow H, Hyman BT, Jack Jr CR, Kawas CH, et al. The diagnosis of dementia due to Alzheimer’s disease: recommendations from the National Institute on Aging-Alzheimer’s Association workgroups on diagnostic guidelines for Alzheimer’s disease. Alzheimer’s & Dementia 2011;7:263–9.

[7] Jack Jr CR, Bennett DA, Blennow K, Carrillo MC, Feldman HH, Frisoni GB, et al. A/T/N: An unbiased descriptive classification scheme for Alzheimer disease biomarkers. Neurology 2016;87:539–47.

[8] Albert MS, DeKosky ST, Dickson D, Dubois B, Feldman HH, Fox NC, et al. The diagnosis of mild cognitive impairment due to Alzheimer’s disease: recommendations from the National Institute on Aging-Alzheimer’s Association workgroups on diagnostic guidelines for Alzheimer’s disease. Alzheimer’s & Dementia 2011;7:270–9.

[9] Birkenbihl C, Salimi Y, Fröhlich H, Initiative JADN, Initiative ADN. Unraveling the heterogeneity in Alzheimer’s disease progression across multiple cohorts and the implications for data-driven disease modeling. Alzheimer’s & Dementia 2022;18:251–61.

[10] Clark LR, Delano-Wood L, Libon DJ, McDonald CR, Nation DA, Bangen KJ, et al. Are empirically-derived subtypes of mild cognitive impairment consistent with conventional subtypes? Journal of the International Neuropsychological Society 2013;19:635–45.

[11] Edmonds EC, Delano-Wood L, Clark LR, Jak AJ, Nation DA, McDonald CR, et al. Susceptibility of the conventional criteria for mild cognitive impairment to false-positive diagnostic errors. Alzheimer’s & Dementia 2015;11:415–24.

[12] Bateman RJ, Benzinger TL, Berry S, Clifford DB, Duggan C, Fagan AM, et al. The DIAN-TU Next Generation Alzheimer’s prevention trial: adaptive design and disease progression model. Alzheimer’s & Dementia 2017;13:8–19.

[13] Levy S-A, Smith G, De Wit L, DeFeis B, Ying G, Amofa P, et al. Behavioral interventions in mild cognitive impairment (MCI): Lessons from a multicomponent program. Neurotherapeutics 2022;19:117–31.

[14] Sherman DS, Mauser J, Nuno M, Sherzai D. The efficacy of cognitive intervention in mild cognitive impairment (MCI): a meta-analysis of outcomes on neuropsychological measures. Neuropsychol Rev 2017;27:440–84.

[15] Jack Jr CR, Bennett DA, Blennow K, Carrillo MC, Dunn B, Haeberlein SB, et al. NIA-AA research framework: toward a biological definition of Alzheimer’s disease. Alzheimer’s & Dementia 2018;14:535–62.

[16] Joshi AD, Pontecorvo MJ, Clark CM, Carpenter AP, Jennings DL, Sadowsky CH, et al. Performance characteristics of amyloid PET with florbetapir F 18 in patients with Alzheimer’s disease and cognitively normal subjects. Journal of Nuclear Medicine 2012;53:378–84.

[17] Chen G, Xu T, Yan Y, Zhou Y, Jiang Y, Melcher K, et al. Amyloid beta: structure, biology and structure-based therapeutic development. Acta Pharmacol Sin 2017;38:1205–35.

[18] Metaxas A, Kempf SJ. Neurofibrillary tangles in Alzheimer’s disease: elucidation of the molecular mechanism by immunohistochemistry and tau protein phospho-proteomics. Neural Regen Res 2016;11:1579–81.

[19] Jia J, Ning Y, Chen M, Wang S, Yang H, Li F, et al. Biomarker changes during 20 years preceding Alzheimer’s disease. New England Journal of Medicine 2024;390:712–22.

[20] Bucci M, Chiotis K, Nordberg A. Alzheimer’s disease profiled by fluid and imaging markers: Tau PET best predicts cognitive decline. Mol Psychiatry 2021;26:5888–98.

[21] Guo Y, Li H-Q, Tan L, Chen S-D, Yang Y-X, Ma Y-H, et al. Discordant Alzheimer’s neurodegenerative biomarkers and their clinical outcomes. Ann Clin Transl Neurol 2020;7:1996–2009.

[22] Ingala S, De Boer C, Masselink LA, Vergari I, Lorenzini L, Blennow K, et al. Application of the ATN classification scheme in a population without dementia: findings from the EPAD cohort. Alzheimer’s & Dementia 2021;17:1189–204.

[23] Van Der Flier WM, Scheltens P. The ATN framework—moving preclinical Alzheimer disease to clinical relevance. JAMA Neurol 2022;79:968–70.

[24] Weber CJ, Carrillo MC, Jagust W, Jack Jr CR, Shaw LM, Trojanowski JQ, et al. The worldwide Alzheimer’s disease neuroimaging initiative: ADNI-3 updates and global perspectives. Alzheimer’s & Dementia: Translational Research & Clinical Interventions 2021;7:e12226.

[25] Edmonds EC, McDonald CR, Marshall A, Thomas KR, Eppig J, Weigand AJ, et al. Early versus late MCI: Improved MCI staging using a neuropsychological approach. Alzheimer’s & Dementia 2019;15:699–708.

[26] Ezzati A, Abdulkadir A, Jack Jr CR, Thompson PM, Harvey DJ, Truelove-Hill M, et al. Predictive value of ATN biomarker profiles in estimating disease progression in Alzheimer’s disease dementia. Alzheimer’s & Dementia 2021;17:1855–67.

[27] Kawahara D, Nagata Y. T1-weighted and T2-weighted MRI image synthesis with convolutional generative adversarial networks. Reports of Practical Oncology and Radiotherapy 2021;26:35–42.

[28] Jagust WJ, Koeppe RA, Rabinovici GD, Villemagne VL, Harrison TM, Landau SM, et al. The ADNI PET Core at 20. Alzheimer’s & Dementia 2024;20:7340–9.

[29] Shaw LM, Vanderstichele H, Knapik-Czajka M, Clark CM, Aisen PS, Petersen RC, et al. Cerebrospinal fluid biomarker signature in Alzheimer’s disease neuroimaging initiative subjects. Ann Neurol 2009;65:403–13.

[30] Salimi Y, Domingo-Fernández D, Bobis-Álvarez C, Hofmann-Apitius M, Birkenbihl C. ADataViewer: exploring semantically harmonized Alzheimer’s disease cohort datasets. Alzheimers Res Ther 2022;14:1–12.

[31] Mueller SG, Weiner MW, Thal LJ, Petersen RC, Jack CR, Jagust W, et al. Ways toward an early diagnosis in Alzheimer’s disease: the Alzheimer’s Disease Neuroimaging Initiative (ADNI). Alzheimer’s & Dementia 2005;1:55–66.

[32] Sled JG, Zijdenbos AP, Evans AC. A nonparametric method for automatic correction of intensity nonuniformity in MRI data. IEEE Trans Med Imaging 2002;17:87–97.

[33] Glover GH, Pelc NJ. Method for correcting image distortion due to gradient nonuniformity 1986.

[34] Jenkinson M, Beckmann CF, Behrens TEJ, Woolrich MW, Smith SM. Fsl. Neuroimage 2012;62:782–90.

[35] Gorgolewski K, Burns CD, Madison C, Clark D, Halchenko YO, Waskom ML, et al. Nipype: a flexible, lightweight and extensible neuroimaging data processing framework in python. Front Neuroinform 2011;5:13.

[36] Ossenkoppele R, Rabinovici GD, Smith R, Cho H, Schöll M, Strandberg O, et al. Discriminative accuracy of [18F] flortaucipir positron emission tomography for Alzheimer disease vs other neurodegenerative disorders. JAMA 2018;320:1151– 62.

[37] Royse SK, Minhas DS, Lopresti BJ, Murphy A, Ward T, Koeppe RA, et al. Validation of amyloid PET positivity thresholds in centiloids: a multisite PET study approach. Alzheimers Res Ther 2021;13:99.

[38] Mattsson-Carlgren N, Andersson E, Janelidze S, Ossenkoppele R, Insel P, Strandberg O, et al. Aβ deposition is associated with increases in soluble and phosphorylated tau that precede a positive Tau PET in Alzheimer’s disease. Sci Adv 2020;6:eaaz2387.

[39] Meyer P-F, Binette AP, Gonneaud J, Breitner JCS, Villeneuve S, Investigators A, et al. Characterization of Alzheimer disease biomarker discrepancies using cerebrospinal fluid phosphorylated tau and AV1451 positron emission tomography. JAMA Neurol 2020;77:508–16.

[40] Fischl B. FreeSurfer. Neuroimage 2012;62:774–81.

[41] Jack Jr CR, Wiste HJ, Weigand SD, Knopman DS, Mielke MM, Vemuri P, et al. Different definitions of neurodegeneration produce similar amyloid/neurodegeneration biomarker group findings. Brain 2015;138:3747–59.

[42] Yu J-T, Li J-Q, Suckling J, Feng L, Pan A, Wang Y-J, et al. Frequency and longitudinal clinical outcomes of Alzheimer’s AT (N) biomarker profiles: a longitudinal study. Alzheimer’s & Dementia 2019;15:1208–17.

[43] Jack CR, Wiste HJ, Weigand SD, Therneau TM, Knopman DS, Lowe V, et al. Age-specific and sex-specific prevalence of cerebral β-amyloidosis, tauopathy, and neurodegeneration in cognitively unimpaired individuals aged 50–95 years: a cross-sectional study. Lancet Neurol 2017;16:435–44.

[44] Filip P, Bednarik P, Eberly LE, Moheet A, Svatkova A, Grohn H, et al. Different FreeSurfer versions might generate different statistical outcomes in case–control comparison studies. Neuroradiology 2022:1–9.

[45] Blennow K, Shaw LM, Stomrud E, Mattsson N, Toledo JB, Buck K, et al. Predicting clinical decline and conversion to Alzheimer’s disease or dementia using novel Elecsys Aβ (1–42), pTau and tTau CSF immunoassays. Sci Rep 2019;9:19024.

[46] Alzubaidi L, Zhang J, Humaidi AJ, Al-Dujaili A, Duan Y, Al-Shamma O, et al. Review of deep learning: concepts, CNN architectures, challenges, applications, future directions. J Big Data 2021;8:1–74.

[47] LeCun Y, Bengio Y, Hinton G. Deep learning. Nature 2015;521:436–44.

[48] Mann HB, Whitney DR. On a test of whether one of two random variables is stochastically larger than the other. The Annals of Mathematical Statistics 1947:50–60.

[49] Cox DR. The regression analysis of binary sequences. J R Stat Soc Series B Stat Methodol 1958;20:215–32.

[50] DeLong LN, Mir RF, Fleuriot JD. Neurosymbolic AI for Reasoning Over Knowledge Graphs: A Survey. IEEE Trans Neural Netw Learn Syst 2024:1–21. 10.1109/TNNLS.2024.3420218.

[51] Lahat D, Adali T, Jutten C. Multimodal data fusion: an overview of methods, challenges, and prospects. Proceedings of the IEEE 2015;103:1449–77.

[52] Huang J, Li Z, Chen B, Samel K, Naik M, Song L, et al. Scallop: From probabilistic deductive databases to scalable differentiable reasoning. Adv Neural Inf Process Syst 2021;34:25134–45.

[53] Salimi Y, Domingo-Fernández D, Hofmann-Apitius M, Birkenbihl C, Initiative ADN, Initiative JADN, et al. Data-driven thresholding statistically biases ATN profiling across cohort datasets. J Prev Alzheimers Dis 2024;11:185–95.

[54] Altman DG, Bland JM. Diagnostic tests. 1: Sensitivity and specificity. BMJ: British Medical Journal 1994;308:1552.

[55] Van Rijsbergen CJ. Information Retrieval, 2nd edn. Newton, MA 1979.

[56] Naidu G, Zuva T, Sibanda EM. A review of evaluation metrics in machine learning algorithms. Computer science on-line conference, 2023, p. 15–25.

[57] Ho TK. Random decision forests. Proceedings of 3rd international conference on document analysis and recognition, vol. 1, 1995, p. 278–82.

[58] Contingency Table. The Concise Encyclopedia of Statistics, New York, NY: Springer; 2008. 10.1007/978-0-387-32833-1_77.

[59] Fisher RA. Statistical methods for research workers. Oliver and Boyd; 1928.

[60] Sánchez-Soblechero A, López-Garc\’\ia S, Lage C, Fernández-Matarrubia M, Irure J, López-Hoyos M, et al. Where Should I Draw the Line: PET-Driven, Data-Driven, or Manufacturer Cut-Off? Journal of Alzheimer’s Disease 2024;98:957– 67.

[61] Veitch DP, Weiner MW, Miller M, Aisen PS, Ashford MA, Beckett LA, et al. The Alzheimer’s Disease Neuroimaging Initiative in the era of Alzheimer’s disease treatment: A review of ADNI studies from 2021 to 2022. Alzheimer’s & Dementia 2024;20:652–94.

[62] Wegner P, Balabin H, Ay MC, Bauermeister S, Killin L, Gallacher J, et al. Semantic harmonization of Alzheimer’s disease datasets using AD-Mapper. Journal of Alzheimer’s Disease 2024;99:1409–23.

[63] Lavin A. Neuro-symbolic neurodegenerative disease modeling as probabilistic programmed deep kernels. International Workshop on Health Intelligence, 2021, p. 49–64.

[64] He Y, Wang Z, Zhang Y, Dan T, Chen T, Wu G, et al. NeuroSymAD: A Neuro-Symbolic Framework for Interpretable Alzheimer’s Disease Diagnosis. ArXiv Preprint ArXiv:250300510 2025.

[65] Ryan J, Fransquet P, Wrigglesworth J, Lacaze P. Phenotypic heterogeneity in dementia: a challenge for epidemiology and biomarker studies. Front Public Health 2018;6:181.

[66] Balabin H, Tamm B, Spruyt L, Dusart N, Kabouche I, Eycken E, et al. Natural language processing-based classification of early Alzheimer’s disease from connected speech. Alzheimer’s & Dementia 2025:e14530.

[67] Zhao Z, Alzubaidi L, Zhang J, Duan Y, Gu Y. A comparison review of transfer learning and self-supervised learning: Definitions, applications, advantages and limitations. Expert Syst Appl 2024;242:122807.

