## Appendix for "Encoding Discordance in the Alzheimer’s Disease A/T/N Framework"

#### **Appendix A: Hyperparameter Optimization**

### All hyperparameters for the Neurosymodal models and the three baseline models were optimized using Bayesian hyperparameter optimization [1,2]. Baseline models were optimized over 500 trials, whereas the Neurosymodal models were optimized over 50 trials due to a higher computational demand. The details for hyperparameter optimization are included in Table A.1 for the Neurosymodal models, Table A.2 for the Random Forest model, and Table A.3 for the logistic regression model. For the baseline probabilistic logic program, the only optimized hyperparameters were the rule probabilities, which were all selected within the range of [0*.*0 - 1*.*0] (Table A.4). For all five models, the AD-specific *F1 score* on the disjoint validation set was optimized. F1 score is a metric which accounts for both false positive and false negative predictions [3].

| **Hyperparameter** | **Range** | **Neurosymodal A Value** | **Neurosymodal B Value** |
| --- | --- | --- | --- |
| **Learning rate** | 1x10^-4^ - 1x10^-3^ | 3.3x10^-4^ | 2.6x10^-4^ |
| **Training batch size** | 4 - 16 | 4 | 12 |
| **Testing batch size** | 4 - 16 | 12 | 12 |

### **Table A.1.** Hyperparameter optimization details for the Neurosymodal models.

| **Hyperparameter** | **Range** | **Selected Value** |
| --- | --- | --- |
| **Number of trees / estimators** | 50 - 300 | 50 |
| **Maximum tree depth** | 2 - 3 | 3 |
| **Minimum samples to split a node** | 2 - 5 | 4 |
| **Minimum samples for a leaf node** | 1 - 10 | 2 |

### **Table A.2.** Hyperparameter optimization details for the Random Forest model.

| **Hyperparameter** | **Range** | **Selected Value** |
| --- | --- | --- |
| **Inverse regularization strength** | 1x10^-4^ - 1.0 | 5.45x10^-2^ |
| **Solver for optimization** | {liblinear, saga} | saga |
| **Penalty type** | {L1, L2} | L1 |

### **Table A.3.** Hyperparameter optimization details for the Logistic Regression model.

| **A/T/N**  **Category** | hasAD(*participant*) | isCU(*participant*) | **Rule Body** | **A/T/N Profile** |
| --- | --- | --- | --- | --- |
| Normal biomarkers | 0.16 | 0.84 | A-(*participant*) ∧T-(*participant*) ∧N-(*participant*) | A−/T−/N− |
| AD continuum | 0.33 | 0.67 | A+(*participant*) ∧T-(*participant*) ∧N-(*participant*) | A+/T−/N− |
|  | 0.86 | 0.14 | A+(*participant*) ∧T+(*participant*) ∧N-(*participant*) | A+/T+/N− |
|  | 0.73 | 0.27 | A+(*participant*) ∧T+(*participant*) ∧N+(*participant*) | A+/T+/N+ |
|  | 0.59 | 0.41 | A+(*participant*) ∧T-(*participant*) ∧N+(*participant*) | A+/T−/N+ |
| Non-AD  pathology | 0.03 | 0.97 | A-(*participant*) ∧T+(*participant*) ∧N-(*participant*) | A−/T+/N− |
|  | 0.28 | 0.78 | A-(*participant*) ∧T-(*participant*) ∧N+(*participant*) | A−/T−/N+ |
|  | 0.41 | 0.59 | A-(*participant*) ∧T+(*participant*) ∧N+(*participant*) | A−/T+/N+ |

**Table A.4.** Optimal rule probability configuration for the baseline probabilistic logic program.

#### **Appendix B: Architecture of the CNN**

### At each of three convolutional layers, the CNN in the neural module works by sliding a 3x3 pixel kernel, with learned parameters, across the MRI at strides of one pixel. These kernels extract the most important information from the MRIs. Thereafter, rectified linear units (ReLU) are applied to introduce nonlinearity [4], and the information is downsampled using maximum pooling by 2x2 pixels and a stride of two. The convolutional and pooling kernel and stride sizes were chosen based on a previous CNN architecture for the classification of AD [5]. Additionally, smaller kernel sizes, such as these, require fewer learned parameters than larger ones, and they can effectively capture complex features when combined with deep NNs [6,7]. The ReLU activation function was chosen since it is commonly used for CNNs (and deep NNs, in general) [4], but also because it promotes sparsity by reducing activations for uninformative input values to zero. In the case of MRIs, uninformative input could include the image space around the brain since it would not inform the prediction task. Thus, promoting sparsity encourages the model to focus on significant features. Finally, to aggregate features, the network includes two fully connected NN layers, with the first layer being subjected to a ReLU activation function for nonlinearity. The output of the second fully connected layer is subjected to a Sigmoid function, which converts a vector of values to probabilities. Here, the probabilities represent the chance of each participant having neurodegeneration (N+) or not (N-).

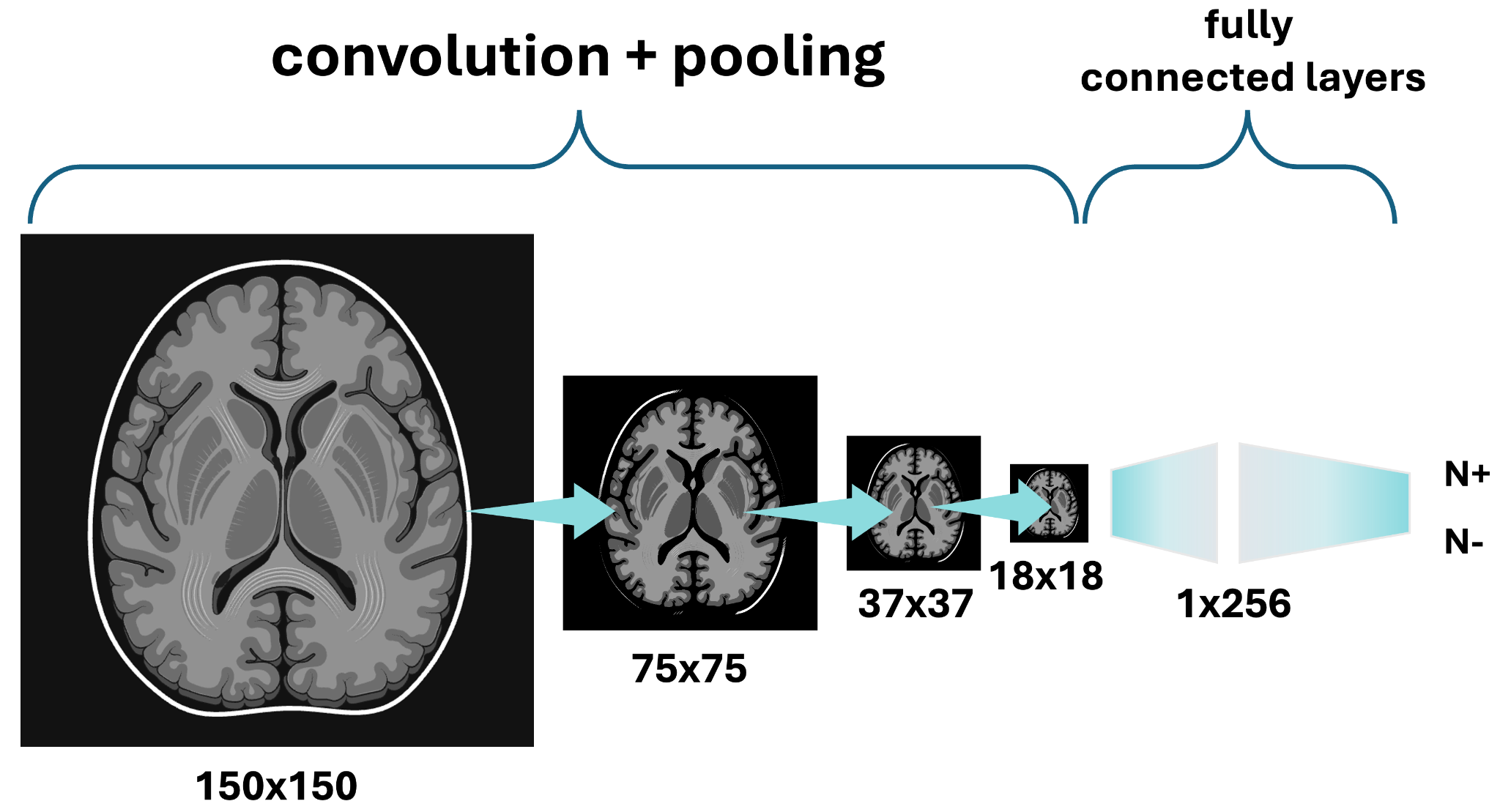

**Figure A.1.** CNN architecture for neurodegeneration determination in both Neurosymodal models. Partially created in https://BioRender.com.

#### **Appendix C: Model Implementation and Training**

### The Neurosymodal Data Fusion pipeline was implemented in an end-to-end manner, so the output of the symbolic module was used to update the parameters of the neural one. This was accomplished through Scallopy, the Python wrapper for Scallop [8]. Scallop was chosen because it is scalable and fully compatible with PyTorch [9], a Python framework for deep learning. Thus, the logic program was written with Scallop and connected to the CNN, which was developed using PyTorch (v. 2.4.1). Learnable rule probabilities were also implemented in PyTorch. The model was trained and executed on one NVIDIA RTX A6000 Graphics Processing Unit [10]. The code to run the Neurosymodal pipeline is packaged and publicly available at [https://github.com/laurendelong21/neurosymodal**.**](https://github.com/laurendelong21/neurosymodal)

### To update model parameters, we computed binary cross entropy loss [11] between true and predicted labels (*i.e.*, CU versus AD). The RAdam optimizer [12] was used for training, which was done in batches of 4 for Neurosymodal model A but 12 for Neurosymodal model B, and learning rates of 3*.*3 × 10^−4^ for 325 epochs and 2*.*6×10^−4^ for 725 epochs were used (for Neurosymodal models A and B, respectively). Training was terminated when the performance on a disjoint validation dataset had plateaued (see Appendix D). Batch sizes and learning rates were optimized as described in Appendix A.

**Appendix D: Early Stopping**

Training was terminated when the running loss on the validation set plateaued. This is otherwise known as early stopping. The running validation loss was computed as the total loss over batched samples in the validation set. Both Neurosymodal models used a batch size of 12 for the validation set. Performance was assessed on the validation set every 25 training epochs, and early stopping was activated after seeing no decrease in validation loss for three consecutive assessments (75 epochs). Running validation losses are shown in Figure A.1 for Neurosymodal model A and Figure A.2 for Neurosymodal model B.

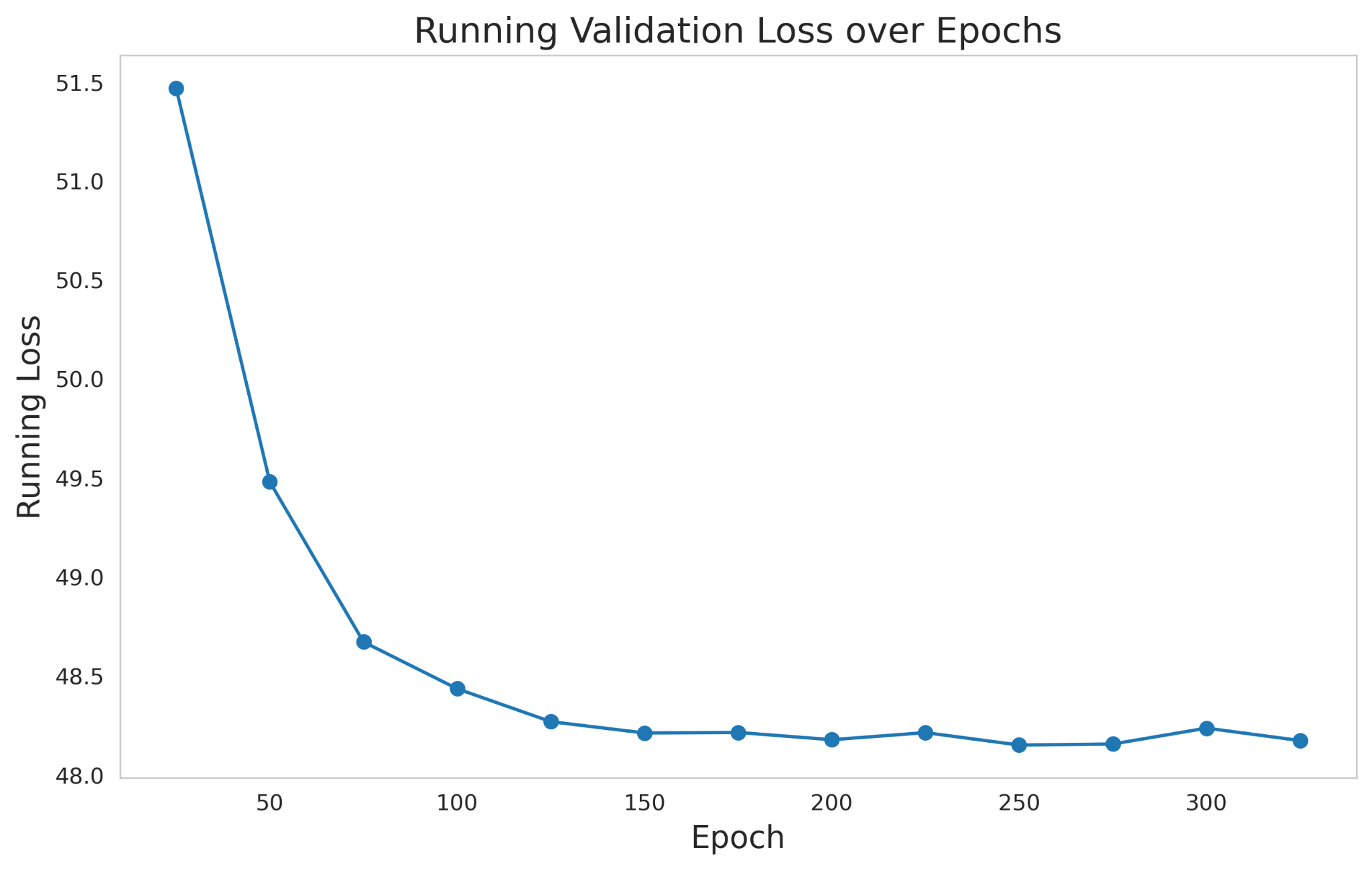
**Figure D.1.** Running validation set losses over 325 epochs of training Neurosymodal model A.
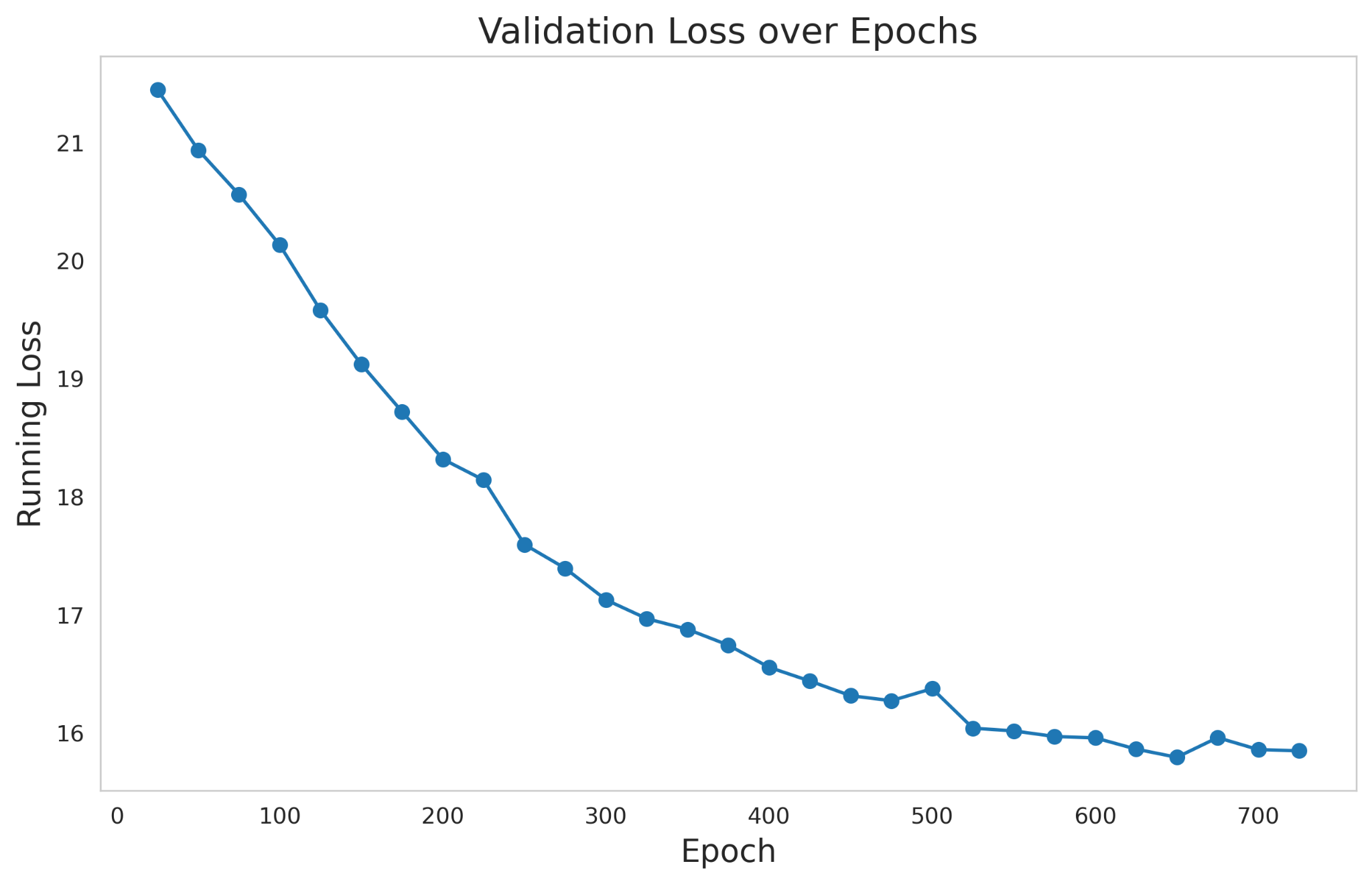
**Figure D.2.** Running validation set losses over 725 epochs of training Neurosymodal model B.

**References**

[1]	Wu J, Chen X-Y, Zhang H, Xiong L-D, Lei H, Deng S-H. Hyperparameter optimization for machine learning models based on Bayesian optimization. Journal of Electronic Science and Technology 2019;17:26–40.

[2]	Bernardo JM, Bayarri MJ, Berger JO, Dawid AP, Heckerman D, Smith AFM, et al. The variational Bayesian EM algorithm for incomplete data: with application to scoring graphical model structures. Bayesian Statistics 2003;7:210.

[3]	Naidu G, Zuva T, Sibanda EM. A review of evaluation metrics in machine learning algorithms. Computer science on-line conference, 2023, p. 15–25.

[4]	Ide H, Kurita T. Improvement of learning for CNN with ReLU activation by sparse regularization. 2017 international joint conference on neural networks (IJCNN), 2017, p. 2684–91.

[5]	Liu M, Li F, Yan H, Wang K, Ma Y, Shen L, et al. A multi-model deep convolutional neural network for automatic hippocampus segmentation and classification in Alzheimer’s disease. Neuroimage 2020;208:116459.

[6]	Naseri H, Mehrdad V. Novel CNN with investigation on accuracy by modifying stride, padding, kernel size and filter numbers. Multimed Tools Appl 2023;82:23673–91.

[7]	Mzoughi H, Njeh I, Wali A, Slima M Ben, BenHamida A, Mhiri C, et al. Deep multi-scale 3D convolutional neural network (CNN) for MRI gliomas brain tumor classification. J Digit Imaging 2020;33:903–15.

[8]	Huang J, Li Z, Chen B, Samel K, Naik M, Song L, et al. Scallop: From probabilistic deductive databases to scalable differentiable reasoning. Adv Neural Inf Process Syst 2021;34:25134–45.

[9]	Imambi S, Prakash KB, Kanagachidambaresan GR. PyTorch. Programming with TensorFlow: Solution for Edge Computing Applications 2021:87–104.

[10]	NVIDIA Corporation. NVIDIA RTX Data Center GPU 2020.

[11]	Zhang Z, Sabuncu M. Generalized cross entropy loss for training deep neural networks with noisy labels. Adv Neural Inf Process Syst 2018;31.

[12]	Liu L, Jiang H, He P, Chen W, Liu X, Gao J, et al. On the Variance of the Adaptive Learning Rate and Beyond. Proceedings of the Eighth International Conference on Learning Representations (ICLR 2020), 2020.
