## Supplementary Material for "Encoding Discordance in the Alzheimer’s Disease A/T/N Framework"

| Criterion | Biomarker | Abbreviation | Cut-off | Source |
| --- | --- | --- | --- | --- |
| A+ | Amyloid-*β*1-42 | A*β* 1-42 | ≤880 pg/mL | CSF |
|  | ^18^F-florbetapir PET SUVR | FBP | ≥1.11 SUVR | PET |
|  | ^18^F-florbetaben PET SUVR | FBB | ≥1.08 SUVR | PET |
| T+ | Phosphorylated Tau 181 | pTau | ≥26.64 pg/mL | CSF |
|  | Temporal meta- ROI PET SUVR | T1 ROI | ≥1.37 SUVR | PET |
|  | Temporal cortex PET SUVR | T2 ROI | ≥1.31 SUVR | PET |
|  | Entorhinal PET SUVR | T3 ROI | ≥1.39 SUVR | PET |
| N+ | Total Tau | tTau | ≥ 300 pg/mL | CSF |
|  | Hippocampal Volume | HVa | *<* 6723 mm^3^ | MRI (FreeSurfer) |
|  | Cortical Thickness | JCT | *<* 2*.*67 mm | MRI (FreeSurfer) |

**Table S1.** Biomarkers for each A/T/N criterion and their respective cut-off values. The cut-off units are specified as follows: pg/mL=picograms per milliliter; SUVR=standardized uptake value ratio; mm=millimeters; mm^3^ =cubic millimeters.

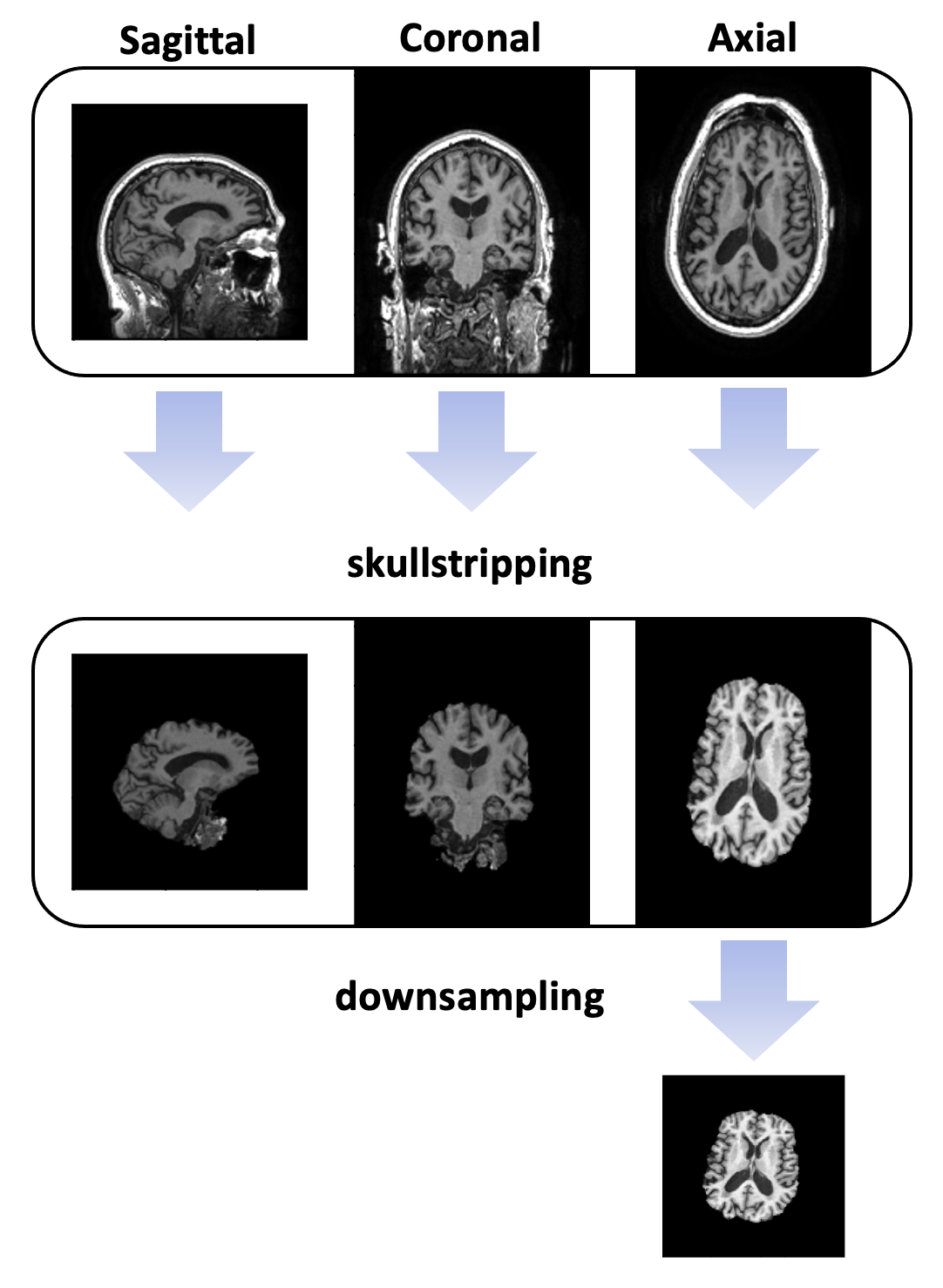

**Figure S1.** MRI processing steps. Each MRI was skullstripped. Then, the center, axial slice was taken and downsampled to 150x150.

| Source | Biomarker | File Name |
| --- | --- | --- |
| CSF | Amyloid-*β*1 − 42 | UPENNBIOMK MASTER FINAL.csv |
| CSF | Phosphorylated Tau 181 (pTau) | UPENNBIOMK MASTER FINAL.csv |
| CSF | Total Tau (tTau) | UPENNBIOMK MASTER FINAL.csv |
| PET | ^18^F-florbetapir (FBP) PET SUVR | UCBERKELEYAV45 8mm 02 17 23.csv |
| PET | ^18^F-florbetaben (FBB) PET SUVR | UCBERKELEYFBB 8mm 02 17 23.csv |
| PET | Temporal metA– ROI (T1) PET SUVR | UCBERKELEYAV1451 8mm 02 17 23.csv |
| PET | Temporal cortex (T2) PET SUVR | UCBERKELEYAV1451 8mm 02 17 23.csv |
| PET | Entorhinal (T3) PET SUVR | UCBERKELEYAV1451 8mm 02 17 23.csv |
| MRI | HVa & JCT | ADNIMERGE download date.csv |
| MRI | HVa & JCT (FreeSurfer 4.3) | UCSFFSX 11 02 15 {download date}.csv |
| MRI | HVa & JCT (FreeSurfer 5.1) | UCSFFSX51 {download date}.csv |
| MRI | HVa & JCT (FreeSurfer 6.0) | UCSFFSX6 {download date}.csv |

**Table S2.** Files downloaded to obtain each of the A/T/N biomarkers.

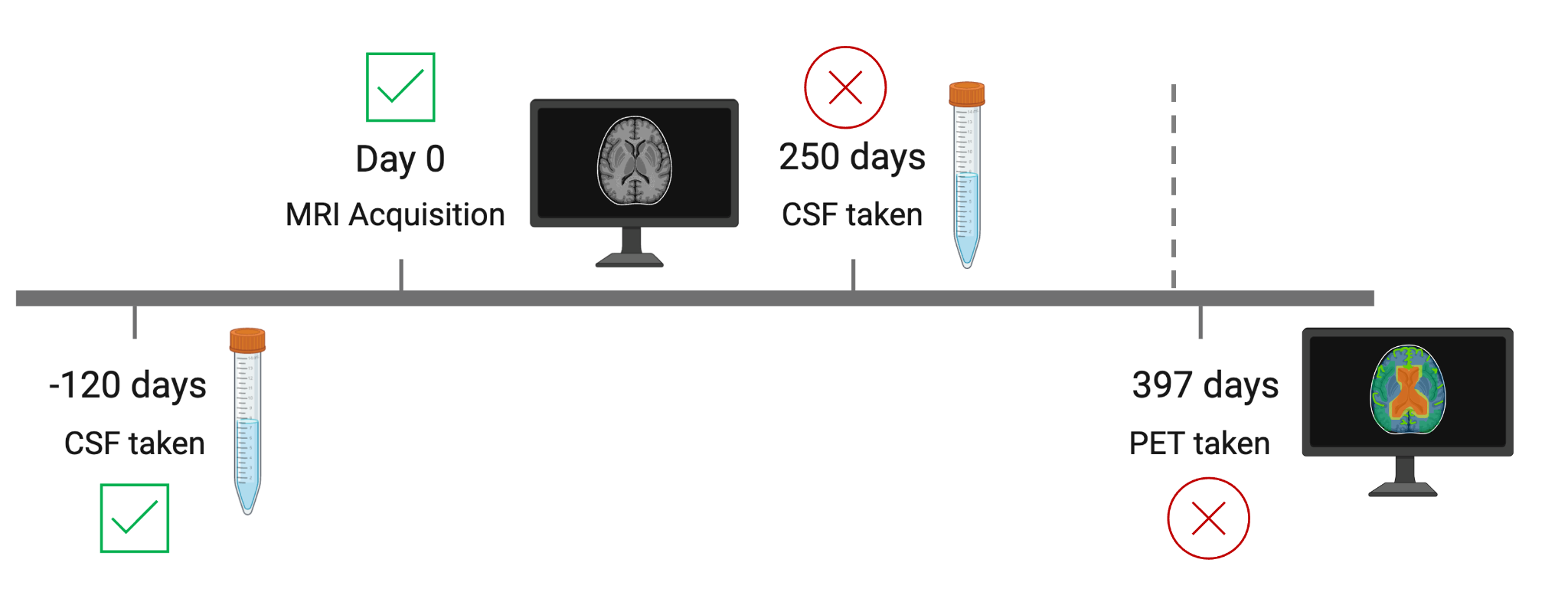

**Figure S2.** **Data point construction.** Each data point comprises an MRI scan and the closest available CSF and PET biomarker information within ±395 days of the MRI acquisition date. In this example, information from the CSF sample taken 120 days prior to MRI acquisition takes precedence over that from the CSF sample taken 250 days after acquisition. Furthermore, the PET data is not be taken into account for this data point since it was taken 397 days after MRI acquisition, so it is not within the specified timeframe. Timeline is not to-scale. Partially created in [https://BioRender.com](https://biorender.com).

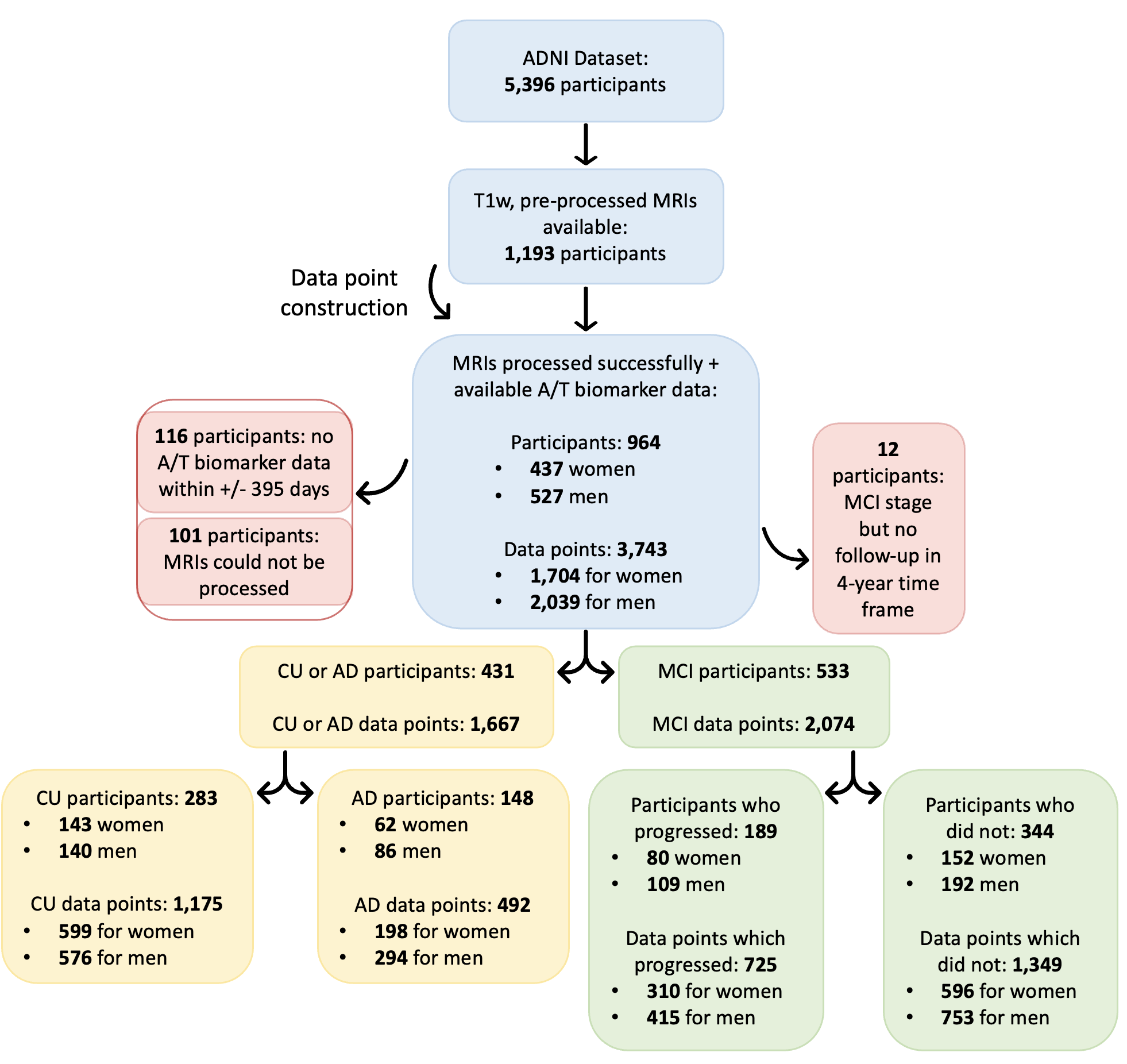

**Figure S3.** **Participant exclusion.**	Flow diagram showing participant and data point inclusion/exclusion.

| **Criterion** | **Num. data points (%)** | **Num. participants (%)** |
| --- | --- | --- |
| **A** | 3,737 (99.9%) | 964 (100.0%) |
| **T** | 2,972 (79.4%) | 809 (83.9%) |
| **N** | 3,350 (89.4%) | 942 (97.7%) |
| **Total** | 3,743 | 964 |

### **Table S3.** **A/T/N Biomarker Availability by Criterion**. Biomarker availability for 3,743 data points. Availability was determined by whether the biomarker collection date was within ±395 days of MRI acquisition for each respective data point. As exceptions, HVa and JCT (N+ biomarkers) were considered available if the FreeSurfer-derived values [18] were available for the respective MRIs.

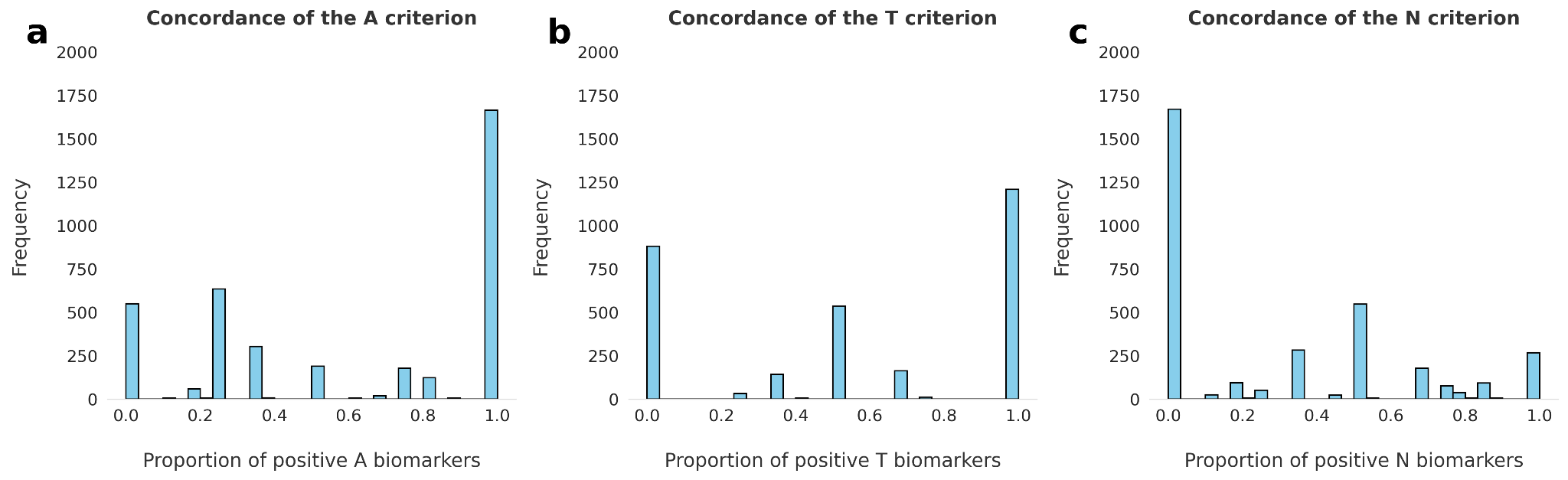

**Figure S4. A/T/N Biomarker Concordances.** Distributions of the proportions of positive **a** A, **b** T, and **c** N biomarkers by data point.

| **Predicate** | **Interpretation** |
| --- | --- |
| A+(*participant*) | participant’s biomarkers show A positivity |
| T+(*participant*) | participant’s biomarkers show T positivity |
| N+(*participant*) | participant’s biomarkers show N positivity |
| hasAD(*participant*) | participant has AD |
| isCU(*participant*) | participant is CU |

**Table S4.** Interpretations of predicates in the probabilistic logic program of module 2.

| **A/T/N**  **Category** | **A/T/N Profile** | **Syndromal Stage** | **Logical Rule** |
| --- | --- | --- | --- |
| Normal biomarkers | A−/T−/N− | AD  CU | hasAD(*participant*) ⇐A-(*participant*) ∧T-(*participant*) ∧N-(*participant*)  isCU(*participant*) ⇐A-(*participant*) ∧T-(*participant*) ∧N-(*participant*) |
| AD continuum | A+/T−/N− | AD  CU | hasAD(*participant*) ⇐A+(*participant*) ∧T-(*participant*) ∧N-(*participant*)  isCU(*participant*) ⇐A+(*participant*) ∧T-(*participant*) ∧N-(*participant*) |
|  | A+/T+/N− | AD  CU | hasAD(*participant*) ⇐A+(*participant*) ∧T+(*participant*) ∧N-(*participant*)  isCU(*participant*) ⇐A+(*participant*) ∧T+(*participant*) ∧N-(*participant*) |
|  | A+/T+/N+ | AD  CU | hasAD(*participant*) ⇐A+(*participant*) ∧T+(*participant*) ∧N+(*participant*)  isCU(*participant*) ⇐A+(*participant*) ∧T+(*participant*) ∧N+(*participant)* |
|  | A+/T−/N+ | AD  CU | hasAD(*participant*) ⇐A+(*participant*) ∧T-(*participant*) ∧N+(*participant*)  isCU(*participant*) ⇐A+(*participant*) ∧T-(*participant*) ∧N+(*participant*) |
| Non-AD  pathology | A−/T+/N− | AD  CU | hasAD(*participant*) ⇐A-(*participant*) ∧T+(*participant*) ∧N-(*participant*)  isCU(*participant*) ⇐A-(*participant*) ∧T+(*participant*) ∧N-(*participant*) |
|  | A−/T−/N+ | AD  CU | hasAD(*participant*) ⇐A-(*participant*) ∧T-(*participant*) ∧N+(*participant*)  isCU(*participant*) ⇐A-(*participant*) ∧T-(*participant*) ∧N+(*participant*) |
|  | A−/T+/N+ | AD  CU | hasAD(*participant*) ⇐A-(*participant*) ∧T+(*participant*) ∧N+(*participant*)  isCU(*participant*) ⇐A-(*participant*) ∧T+(*participant*) ∧N+(*participant*) |

**Table S5.** Logical rules encoding all A/T/N profiles as rule bodies and CU and AD syndromal stages as rule heads.
